# CLINICOPATHOLOGICAL AND MOLECULAR CHARACTERISTICS OF PLASMA CELL RICH REJECTION IN RENAL TRANSPLANT BIOPSIES

**DOI:** 10.1101/2024.06.27.24309579

**Authors:** Romy du Long, Marie S.N. Chevalier Florquin, Frederike J. Bemelman, Nike Claessen, Hessel Peters-Sengers, Sandrine Florquin, Jesper Kers

## Abstract

**Background:** Plasma cell rich rejection (PCRR) is an uncommon, ill-defined type of renal allograft rejection in the current literature considered a subtype of T cell-mediated rejection (TCMR). PCRR has poorer clinical outcome and is often refractory to classic immunosuppressive therapy. Our study analyzed clinical course, Banff lesion scores and mRNA expression of PCRR compared to (late) rejection.

**Methods:** We retrospectively scored and reclassified the last known biopsy of 263 renal transplant recipients, morphologically classified as rejection according to the 2019 Banff classification. mRNA expression analysis was performed using the Nanostring B-HOT panel on a subset of cases. PCRR was compared to (late) TCMR, ABMR and mixed rejection for renal function follow-up and graft survival.

**Results:** mRNA analysis revealed uniquely expressed genes in PCRR including *LOX*, *CPA3*, *IL4*, *IL17F,* and *MMP12*. PCRR is enriched for genes related to mast cells, memory B- and T-cells and transcripts involved in NK cells and allograft fibrosis with heterogeneity in gene expression in biopsies with PCRR. PCRR might be a late event compared to late TCMR and ABMR, with a higher degree of total inflammation and fibrosis. Graft survival and renal function was similar to late TCMR and ABMR during a 5-year follow-up period after renal biopsy.

**Conclusion:** PCRR represents a distinct late-onset stage of inflammation displaying diverse gene expression patterns, with presence of mainly mast cells, NK cells and transcripts involved in renal allograft fibrosis. Clinical outcomes in patients with PCRR appeared more similar to late TCMR and ABMR.

## INTRODUCTION

Renal transplantation is the primary therapy of choice for patients afflicted by end-stage kidney disease (ESKD). Despite increasing success of current immunosuppression, allograft rejection is still a significant post-transplant complication leading to graft loss. Allograft rejection is categorized into T cell-mediated rejection (TCMR), antibody-mediated rejection (ABMR), or a combination of both termed mixed rejection. A morphologically distinct presentation of allograft biopsies is plasma cell rich rejection (PCRR), but this is not considered a separate type of rejection in the current Banff classification for kidney allograft.^1^ PCRR has previously been described in liver transplants and referenced within the Banff schema for liver allograft rejection.^2^ PCRR in liver grafts was designated *de novo* autoimmune/plasma cell hepatitis due to its resemblance to native liver autoimmune hepatitis (AIH) and overlapping etiology with autoimmunity, and has been identified as a causative factor for late graft dysfunction. PCRR still remains an ill-defined form of renal allograft rejection due to the current lack of knowledge about its pathogenesis and molecular mechanisms. Histologically, PCRR is characterized by the presence of plasma cells, either diffusely distributed in the renal tissue or aggregated into clusters, often accompanied by myxoid interstitial edema, with variable degrees of tubulitis, arteritis, microvascular inflammation and often capillary C4d deposition with or without donor-specific antibodies.^2–4^

In general, PCRR manifests at a later time post-transplant than TCMR, ranging from 302 days to 3.7 years post-transplant according to current literature.^3–7^ The majority of patients with PCRR present with histopathological patterns similar to TMCR type I or II.^8^ As a result, PCRR is considered a subtype of TMCR rather than a distinct entity. However, recent studies have highlighted various dissimilarities between PCRR and ‘classic’ TMCR concerning molecular and clinic-pathologic features, supporting the hypothesis that PCRR could be considered a distinct type of allograft rejection. One hypothesis states PCRR is mainly associated with a combination of inadequate immunosuppression in conjunction with preceding viral infections.^9^ Another hypothesis poses that PCRR is an exacerbated presentation of chronic rejection by persistent activation of the immune system and generation of antibody-secreting plasma cells, either generated *in situ* or recruited to the graft.^10^

Based on previous research it has been suggested PCRR is associated with a poorer clinical outcome compared to TMCR and ABMR, including a more rapid decline of renal function and higher rate of renal graft loss.^3,5,10,11^ Nonetheless, some studies have not identified a correlation between the presence of plasma cells and an adverse clinical outcome.^12,13^ In clinical practice, PCRR is more often refractory to classical immunotherapy regimes, thereby complicating the standardized management of this type of rejection.^6,8^ These differences in clinical progression and graft survival could indicate PCRR is driven by different underlying immunological mechanisms in comparison to TCMR and ABMR. Unraveling its underlying molecular and immunological determinants is key to attain better insights into the cellular pathways and processes involved in this form of rejection.

In this study, we analyzed PCRR in a large cohort of patients who underwent renal transplantation at the Amsterdam UMC. Our objective was to study clinical, histological and molecular signatures of PCRR in comparison to other types of rejection, especially late presentations of TCMR and ABMR. We hypothesized PCRR could be a specific subtype of allograft rejection with different molecular and cellular characteristics compared to (late) TCMR and ABMR with differences in clinical outcome.

## MATERIAL AND METHODS

### Selection of patients and renal biopsies

For the epidemiological study, the archives of the Amsterdam UMC, location AMC, were screened for all kidney transplant biopsies performed between 01-01-2000 and 01-06-2018 (N = 3376 biopsies). The number of biopsies per patient ranged from 1 to 12 (average 1.94/patient). After exclusion of biopsies for external consultation, patients with only an implantation biopsy, biopsies of renal tumor tissue or the lack of stained-glass slides, a total of 1055 biopsies from 1055 individual transplant recipients were included. For a comprehensive explanation of the inclusion criteria and process, see the original article by Kers et. al.^14^ All 1055 biopsies were re-assessed by one expert kidney pathologist (SF) according to the latest update of the Banff classification in 2019 in terms of Banff lesion scores and diagnostic classes.^1^ Slides were analyzed based on the hematoxylin and eosin (H&E), periodic acid-Schiff (PAS) and Jones silver staining. Cases classified as rejection, either (active/chronic) TCMR, (active/chronic) ABMR or mixed rejection as described in the most recent 2019 Banff classification, were included for subsequent comparison. Cases with borderline rejection were excluded from this study. In order to directly compare the association between biopsy findings and graft outcome, and eliminating any possible intervening acute biopsy-triggering events between the moment of biopsy and end of follow-up, the last biopsy of each of the patients with a documented proven episode of rejection was included. For this study, we were also interested in comparing PCRR to a subset of only late type of rejection, and by including the last biopsy it ensured inclusion of biopsies with sufficient chronicity. Histologically, we defined ongoing late type of rejection as chronic-active TCMR (caTCMR), chronic-active ABMR (caABMR) and chronic-active mixed rejection (caMixed), as defined in the most recent Banff 2019 classification. All cases with plasma cell rich infiltrates, defined as infiltrates with more than 10% plasma cells based on a definition used in previous studies, were classified as PCRR and included.^3,7,15^ Concurrent viral infections, e.g., BK nephropathy, were excluded based on immunohistochemical staining (SV40 LTAg). Inadequate biopsies as defined by the Banff classification (minimal of 7 glomeruli and one artery) and patients lacking relevant clinical data were excluded. The final cohort comprised a total of 263 patients/biopsies with complete clinical follow-up data for graft survival and kidney graft function during a follow-up period of at least 3 years after the last included biopsy (**Figure 1**). Extensive clinical data were ascertained from the patient’s electronic hospital files, including recipient characteristics (i.e., age, gender, HLA mismatches, underlying diseases, primary diagnosis), donor characteristics (i.e., age, gender, type of donation), transplantation-related characteristics (cold and warm ischemia time, delayed graft function) and clinical parameters (renal function). The presence of donor specific antibodies (DSA), either preformed of *de novo*, were measured using the Luminex single antigen beads assay.

**Figure 1.**
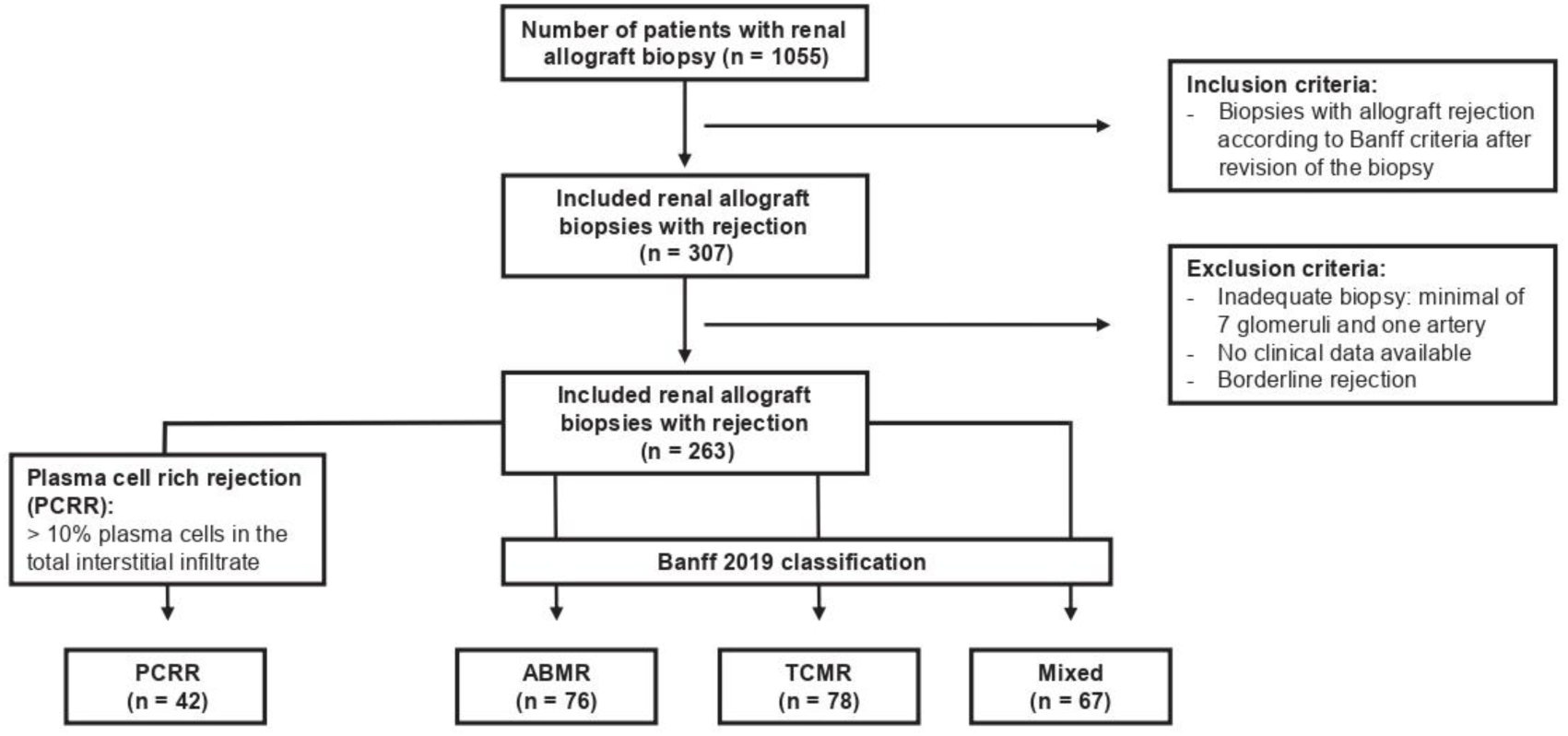
– Selection of patients and renal allograft biopsies. Flowchart of included patients and renal allograft biopsies. Abbreviations: PCRR = plasma cell rich rejection, ABMR = antibody mediated rejection, TCMR = T-cell mediated rejection

### mRNA expression analysis on biopsy tissue

To examine gene expression profiles in biopsies with rejection, we used the Nanostring B-HOT panel specifically designed for organ transplantation. The nCounter^TM^ Human Organ Transplant Panel (B-HOT panel), including 770 genes (of which 10 housekeeping genes), involves 37 pathways including tissue damage, organ rejection and immune responses. ^16^ Several recent studies showed this panel aids the classification of kidney transplant rejection and proved to be feasible from a technical standpoint. ^17,18^ We selected an additional set of 24 biopsies (8 biopsies with ABMR, 8 with TCMR and 8 with PCRR) from renal transplant patients between 01-01-2018 and 31-12-2019 from the Amsterdam UMC. We selected biopsies based on histological criteria for ABMR, TCMR or PCRR without signs of any other histopathological abnormalities after histological revision of the slides by an expert renal pathologist (archetypical cases of rejection phenotypes) (**Supplementary Table 1**). These biopsies were also scored according to the most recent 2019 Banff classification. ^1^ Five whole tissue sections of 20 µm from FFPE blocks were obtained from each biopsy for mRNA isolation. After xylene deparaffinization, mRNA was isolated using the Nucleospin Total RNA FFPE XS Isolation Kit (MN, item number 740969.50). The mRNA quality was assessed with the Agilent TapeStation after which 50 ng of mRNA was incubated for 20 hours overnight. R programming software (version RStudio 2023.12.1+402 “Ocean Storm” Release for Windows 11, www.r-project.org) and nSolver Analysis Software (Nanostring Technologies, version 4.0) were used to analyze and visualize the data. To identify which cell types might play a role in PCRR, we performed CIBERSORT, a method described by Newman et al. used to characterize cell composition in complex human tissue based on their gene expression profiles.^19^ Next, we performed pathway and cell type analysis using predefined gene sets based on extensive research by the group of Halloran et al. who defined pathogenesis-based transcript sets (PBT) to determine the molecular phenotype in renal transplants.^20^ The transcripts were based on previously discovered, defined and validated gene sets based on extensive microarray studies and these gene sets have been shown to correlate with different types of rejection, Banff lesion scores and clinical outcome.^21–25^ The gene sets evaluated in our study included DSAST, DSA10, ENDAT, NK cell, TCMR, Progression GoCAR and Extracellular Matrix Organization.^21,26–29^ (See **Supplementary Table 2**) These gene sets consisted of the same genes as defined in the study by Rosales et al., using the same B-HOT panel in our study for mRNA analysis.^30^ We also added three of the mast cell associated transcripts (MACAT) described by Mengel et al., including *CPA3*, *TPSAB1* and *MS4A2*.^31^

### Rejection Class Algorithm

Recently, a model was built and developed by the KU Leuven with the use of semisupervised and data-driven clustering methods to produce phenotypic reclassification of kidney transplant rejection that is both histologically and clinically relevant.^32,33^ The current Banff classification stratifies renal transplant biopsies into five categories based on a combination of acute and chronic semiquantitative, ordinal lesions scores.^1^ However, these categories are not mutually exclusive and Banff lesions scores are not specific for certain types of rejection, making it more difficult to accurately diagnoses renal transplant biopsies. This model identified six new relevant cluster phenotypes in acute kidney transplant rejection which were significantly associated with graft failure.^33^ Based on this model, new phenotypes and disease classification are defined, as a complementary tool along the Banff classification. Applying the clustering method to chronic histologic lesions, four chronic phenotypes were identified.^32^ The model was translated to a web-based software classed ‘RejectClas’ for research purposes which is freely accessible via their website https://rejectionclass.eu.pythonanywhere.com. We uploaded our data containing the Banff scores for acute lesions and chronic lesions in a format as described on their website. Next, the software stratified the biopsies along the radius axis in different strata, which represents the extent inflammation activity (activity index) and total chronicity (chronicity index). Using this model and software, we evaluated the activity index and chronicity index of all biopsies included in our study. We compared the results from biopsies with PCRR to these with ABMR, TCMR and mixed rejection to determine whether PCRR is similar to these types of rejection based on their activity index and chronicity index. For an extensive explanation of the validation process and clustering methods, see the originally published articles by Vaulet et al.^32,33^

### Immunostainings

After Nanostring analysis additional immunohistochemical stains were performed on a subset of 42 renal biopsies (10 biopsies with PCRR, 12 biopsies with mixed rejection, 10 biopsies with ABMR and 10 biopsies with TCMR). The slides were stained using CD3 for T lymphocytes (anti-rabbit Ab, RM9107 Thermo), CD20 for B lymphocytes (anti-mouse Ab, M0755 DAKO), CD68 for macrophages (anti-mouse Ab, M0876 DAKO), CD138 for plasma cells (anti-mouse Ab, 3825-C1 WRKLM), Foxp3 for T-regulatory cells (anti-rat Ab, 14-4776-82 eBioscience) and cKit for mast cells (anti-rabbit Ab, 117R-16 CellMarque). CD3, CD20 and CD68 were quantified by scoring the percentage of positive cells in the interstitial infiltrate in 10 randomly selected high-power fields (HPF) and calculate the mean percentage of positive cells/mm2. CD138, cKit and Foxp3 were quantified by counting the number of positive cells in 10 randomly selected HPF with interstitial infiltrates and calculating the mean number of cells/mm^2^. For both methods the ImageJ software (version ImageJ 1.53k for Windows) was used.

### Clinical outcomes

The primary clinical outcomes evaluated in this study included death-censored graft failure, defined as return to dialysis or retransplantation, and renal function (as indicated by serum creatinine levels). Secondary clinical outcomes included time between renal transplantation and development of an episode of rejection and development of DSA. In a subanalysis we performed a comparative analysis of the same clinical outcomes as described above in PCRR to a subgroup of patients with only ongoing late types of rejection. This subanalysis aimed to investigate whether clinical outcomes of PCRR demonstrate closer similarity to late rejection, which cause late graft dysfunction with adverse long-term prognosis.^34^

### Statistical Analysis

All other statistical analyses were performed within the R computing environment in the Rstudio software (version RStudio 2023.12.1+402 “Ocean Storm” Release for Windows 11). Categorical outcomes were analyzed using the Chi-square test, while continuous variables were analyzed using the unpaired t-test, non-parametric Mann-Whitney U test or Kruskal-Wallis test depending on the underlying characteristics of the variables and outcome. Pairwise comparisons were performed using Dunn’s procedure with a Bonferroni correction for multiple comparisons. Survival analysis was performed using Kaplan-Meier curves and log-rank test to compare death-censored graft failure between different groups. With a Cox-proportional hazards model hazard ratios (HR) and 95% confidence intervals for death-censored graft failure were calculated. Odds ratio and risk factors for the development of PCRR were identified with univariate regression analysis. First, linear mixed models’ analysis was performed to investigate the longitudinal outcome trajectory of renal function of PCRR in comparison to other types of rejection. Renal function was expressed as serum creatinine (µmol/L) and log_10_ transformed to better approach a normal distribution. Second, a joint models (JMs) for longitudinal and time-to-event data was used to avoid biased estimated of renal function decline.^35^ This model combines a linear mixed model to a Cox proportional hazards model, capturing the risk of death-censored graft failure. The main follow-up period for was a 5-year period starting at the moment of the last included biopsy. To adjust for immortal time bias as a result of differences in time between the moment of renal transplant and the last biopsy a subsequent landmark analysis as described by Gleiss et al. was performed.^36^ In the landmark analysis, a period of time between de baseline time (e.g. moment of renal transplant) and follow-up start date (the landmark date) is chosen a priori and designated as the exposure period. Only patients with a clinical outcome occurring after the landmark date are included in the analysis. Hence, patients who experience the outcome of interest during the exposure period are excluded from the subsequent analysis to avoid reverse causality and immortal time bias, due to the fact the moment of the last included biopsy was not equal in all patients. A sensitivity analysis using a landmark at one and two years after renal transplant were performed. For the assessment of death-censored graft failure and renal function, loss to follow up due to death or graft failure within the exposure window were used as exposure variables. See **Supplementary Methods** for a more extensive description of the statistical analysis.

## RESULTS

### Baseline characteristics

In our study, 42 patients (16%) were diagnosed as PCRR, 76 as ABMR (29%), 78 as TCMR (30%) and 67 as mixed rejection (25%). The prevalence of PCRR in our study was 4% (42/1055), similar to prior research ranging from 2 to 5%.^3,5,7^ The median time from renal transplantation to the episode of rejection in the last included biopsy for the entire cohort was 0.5 year (interquartile range [IQR] 0.0-2.3 years), with the median length of the follow-up period after the last included biopsy 3.0 years (IQR 0.5-7.0 years). Detailed baseline characteristics for each group are presented in **Table 1**.

**Table 1.**
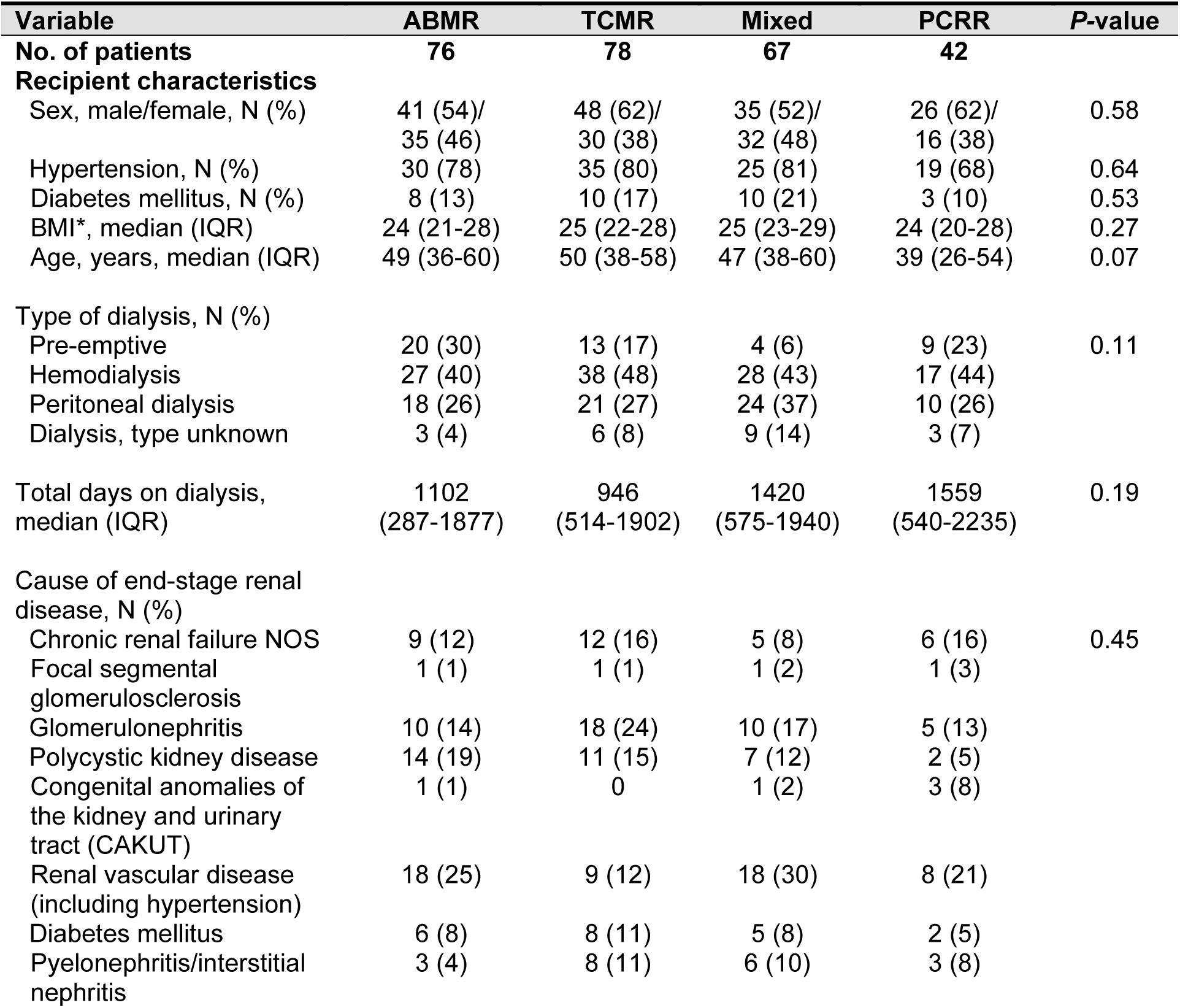
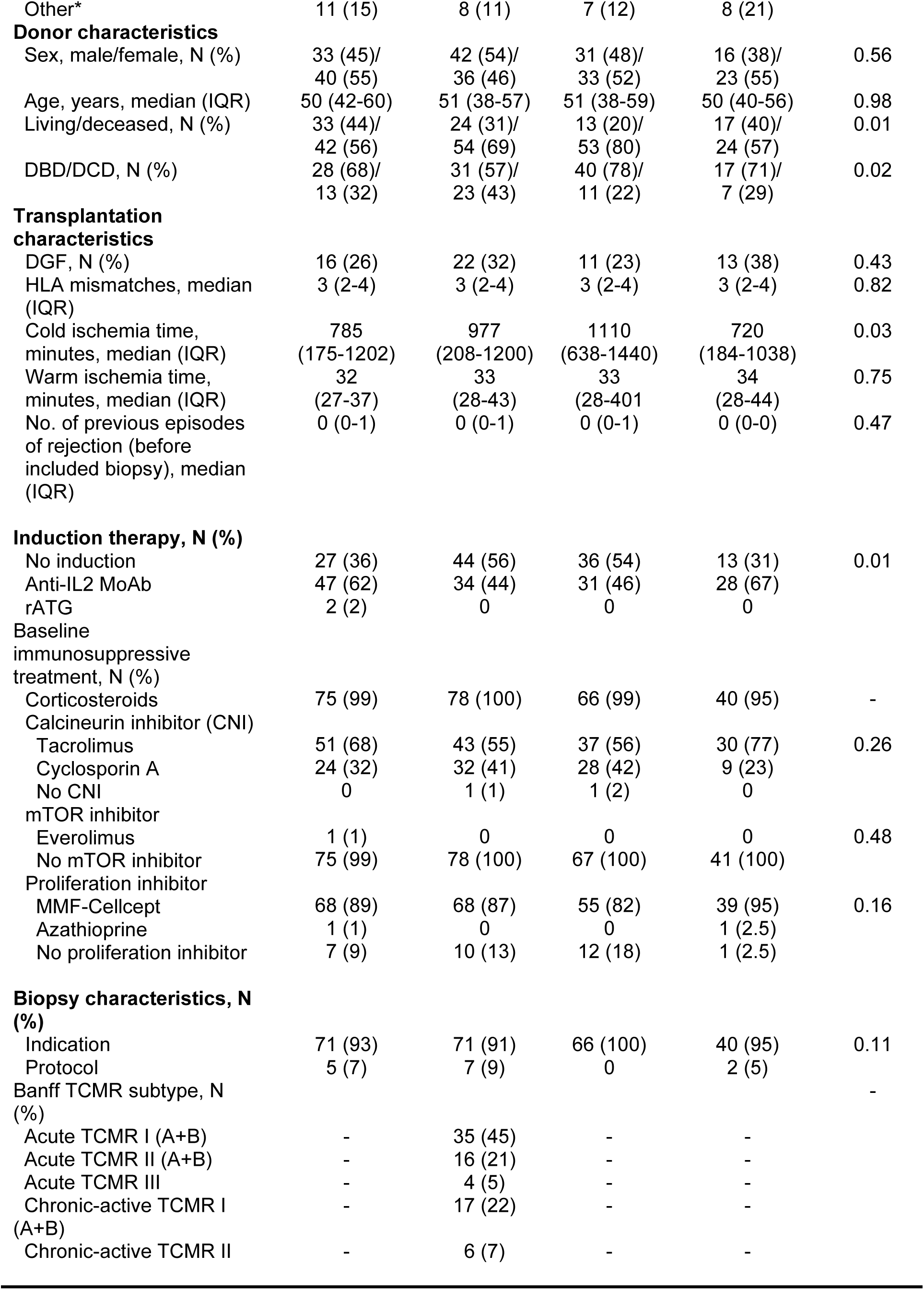
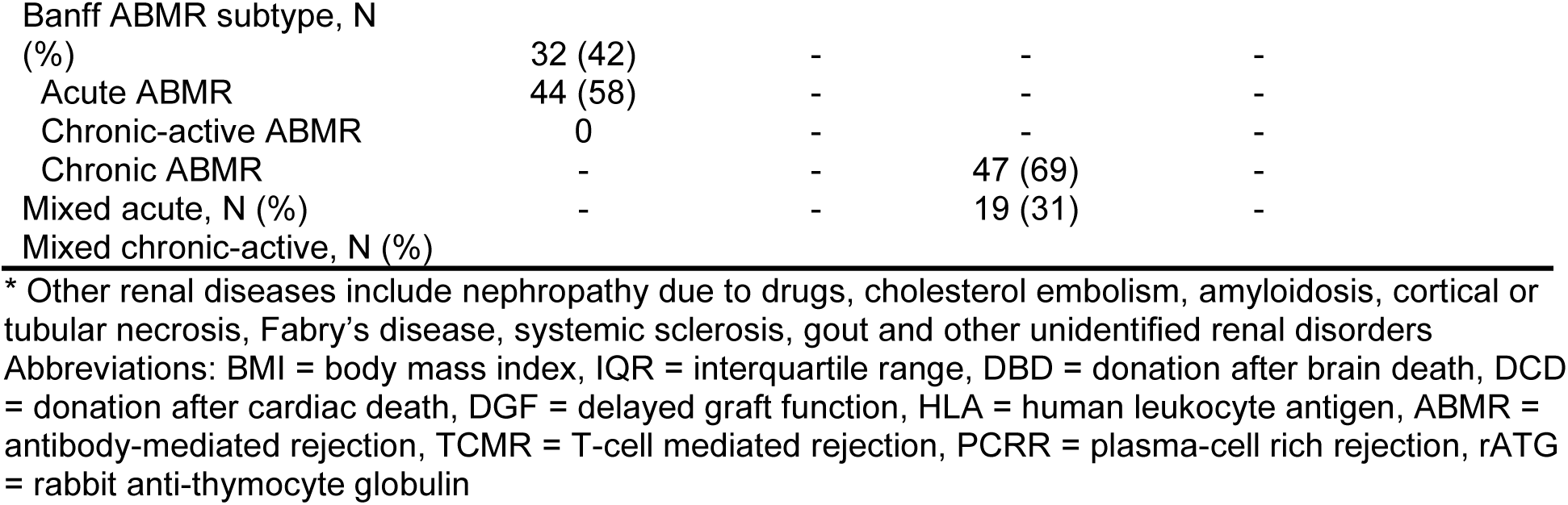
– Baseline clinical and biopsy characteristics of all included patients.

### mRNA expression analysis identifies PCRR as an inflammatory state molecularly distinct from TCMR and ABMR

In PCRR, an upregulation in genes associated with tissue remodeling (*MMP12*, *LOX*), mast cells (*IL4*, *CPA3*) and B cell signaling (*NFMA1*) was observed (**Figure 2A/B**). Various cytokines and growth factors (*TNFSF4*, *IL12RB2* and *IL17F*) exhibited differential expression patterns compared to TCMR and ABMR. In a principal component analysis (PCA), biopsies with PCRR displayed larger heterogeneity in gene expression than TCMR and ABMR (**Figure 2C**). Similar results were observed with stochastic neighborhood embedding (t-SNE) (**Figure 2D**). Next, a PCA based on immunohistochemical quantifications in PCRR biopsies demonstrated more heterogeneous cell types in the infiltrate compared to TCMR and ABMR, possibly reflecting the heterogeneity previously seen in the gene expression patterns (**Supplementary Figure 1**). In a pseudotime trajectory analysis (**Figure 2E/F**) PCRR showed less convergence with TCMR and ABMR. This might imply that PCRR is a delayed manifestation of a similar disease process seen in ABMR and TCMR, or potentially a distinct disease process guided by different underlying gene expression profiles.

**Figure 2.**
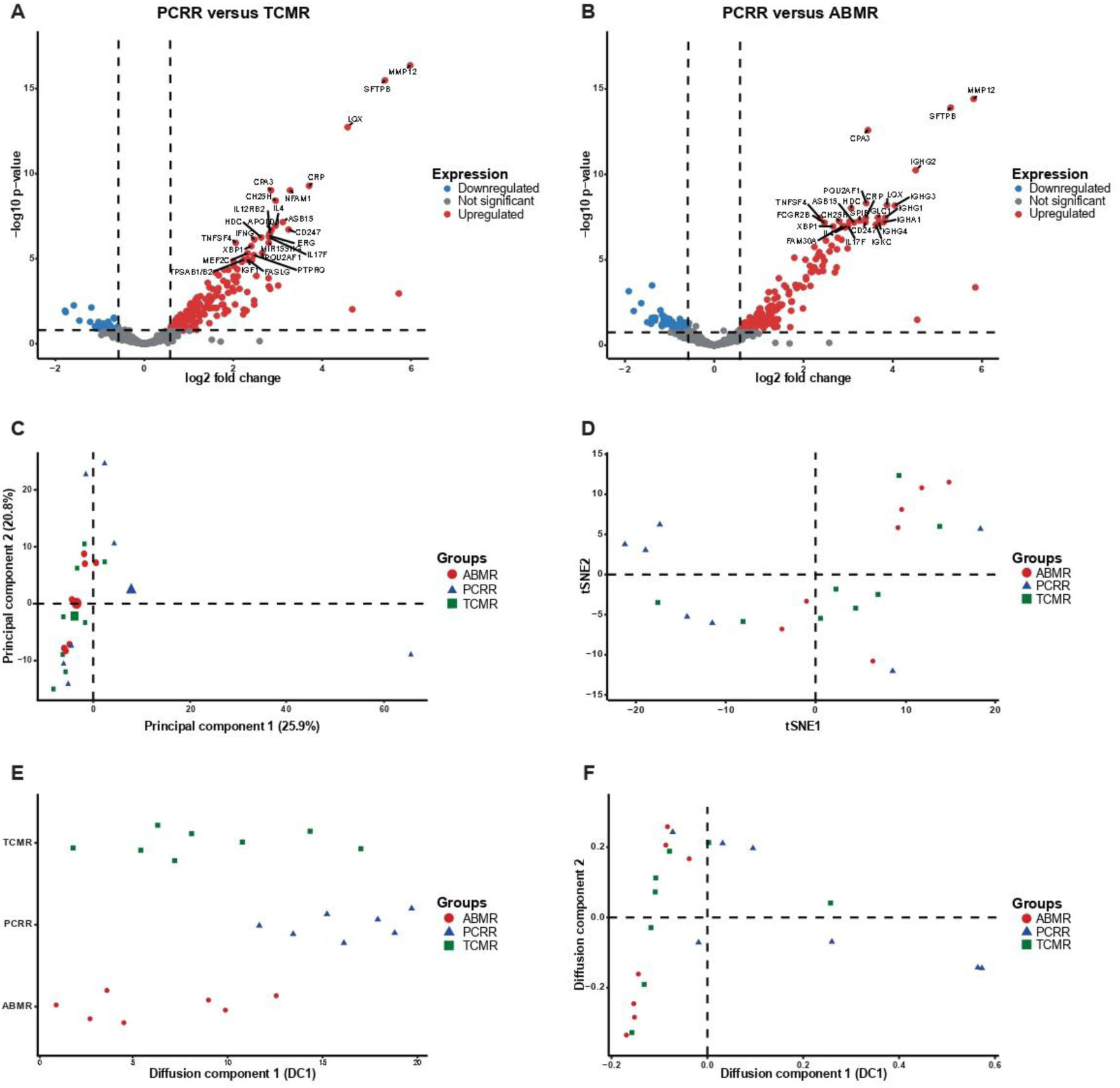
- Nanostring mRNA analysis. Volcano plots showing the most significantly upregulated differentially expressed genes in biopsies with PCRR compared to biopsies with TCMR (**A**) or ABMR (**B**). (**C**) A PCA plot showing differences in gene expression (heterogeneity) within the individual biopsies with PCRR (triangles) compared to only TCMR (rectangles) or ABMR (circles). A tSNE plot (**D**) visualizing the variance in gene expression in PCRR, compared to TCMR and ABMR. Pseudotime analysis (E and F) comparing PCRR to TCMR and ABMR. Abbreviations: ABMR = antibody mediated rejection, TCMR = T-cell mediated rejection, PCRR = plasma cell rich rejection, tSNE = t-distributed Stochastic Neighbor Embedding GSEA identified notable enrichment of plasma cells, NK cells and monocytes/macrophages in PCRR compared to TCMR (**Figure 3A/B**). In comparison to ABMR, PCRR was enriched for mast cells, B cells (naïve and memory) and different CD4+ T helper subsets (resting memory, naive, follicular and regulatory) and CD8+ T cells. NK cell transcripts and Progression GoCAR were the topmost upregulated gene sets in PCRR (**Figure 3C/D**). Additionally, MACAT was upregulated due to the significant enrichment of *CPA3*. An increased number of mast cells was confirmed with immunostainings (PCRR median 47.2 cells/mm^2^ (IQR 39.0-58.5), ABRM 10.6 (IQR 2.6-12.8, *P*=0.001), TCMR 20.3 (IQR 15.2-47.0, *P*=0.20) and mixed rejection 19.5 (IQR 6.9-27.2, *P*=0.03); **Supplementary Figure 2**). There was a strong positive correlation between fibrosis (based on the IF/TA-score) and the number of mast cells in all biopsies combined (*r*(29)=0.32, *P*=0.002). No differences were observed in the other immunostainings (CD3, CD20, CD68 and FoxP3).

**Figure 3.**
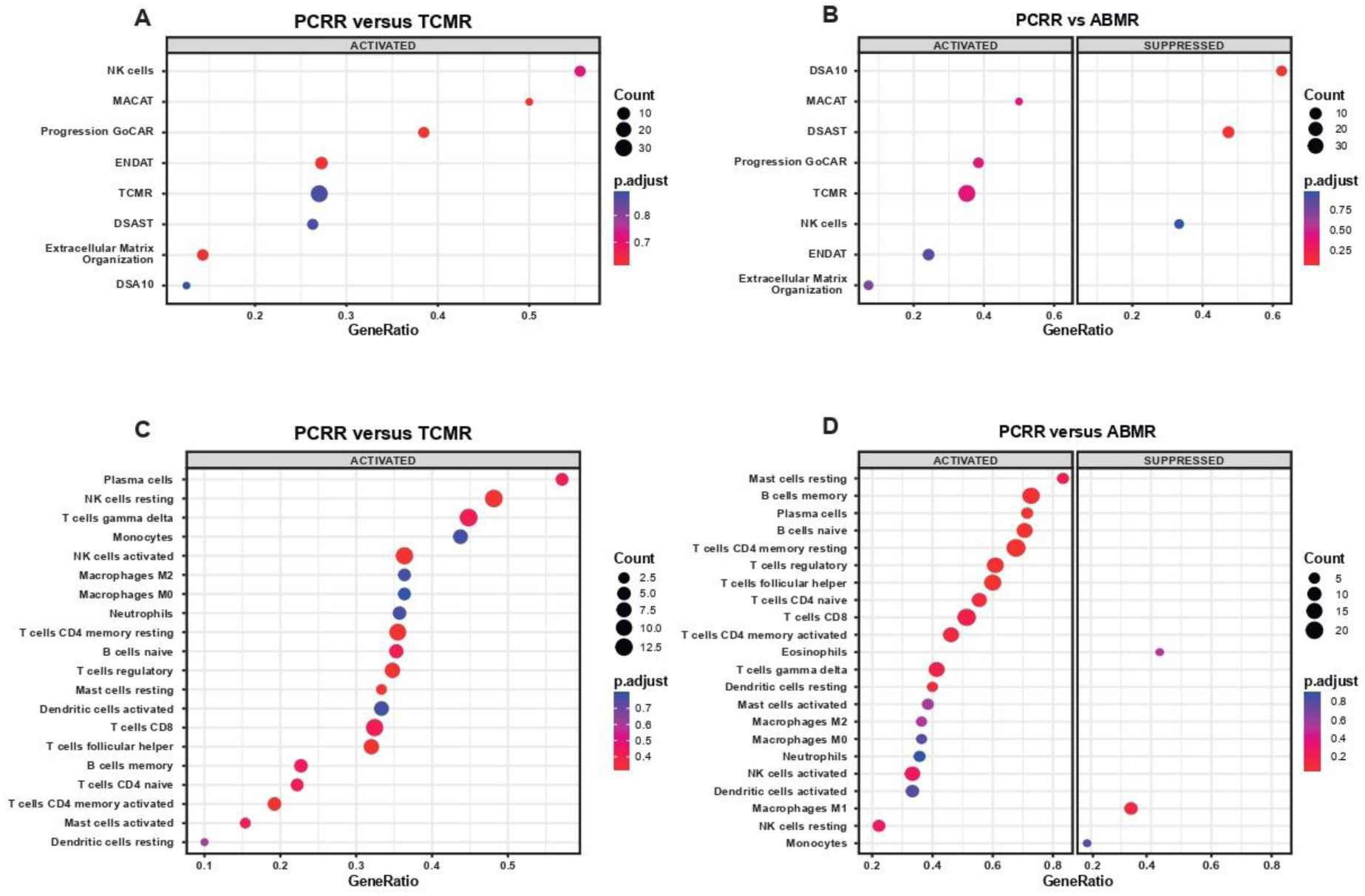
- Gene set enrichment analysis (GSEA) **T**he top two panels showing the upregulated gene sets in biopsies with **(A)** PCRR in comparison to TCMR and **(B)** ABMR. The used gene sets are based on the gene sets as defined in the article of Rosales et al. consisting of genes that are present in the Nanostring B-HOT panel.^29^ The lower two panels show different cell types either upregulated or downregulated in **(C)** PCRR compared to TCMR and **(D)** ABMR. Abbreviations: ABMR = antibody mediated rejection, TCMR = T-cell mediated rejection, PCRR = plasma cell rich rejection, DSAST = donor specific antibody selective transcripts, ENDAT = endothelial associated transcripts, DSA10 = top 10 transcripts associated with donor specific antibodies, NK cells = natural killer cells transcripts, Progression GoCar = Genomics of Chronic Allograft Rejection gene transcripts, TCMR = T-cell mediated rejection selective transcripts, MACAT = mast cell associated transcripts.

### Histological scoring of biopsies (Banff classification)

PCRR displayed more graft inflammation reflected by more tubulitis, interstitial inflammation and total inflammation (**Table 2, Supplementary Table 3**). Furthermore, PCRR exhibited the highest degree of chronic damage (TCMR median IF/TA 20% (IQR 10-30%), ABMR 20% (IQR 10-30%), mixed rejection 10% (IQR 10-30%) and PCRR 40% (IQR 20-70%), *P*=0.001). Compared to late rejection, PCRR continued to exhibit of a high degree of graft inflammation and chronic damage (caTCMR IF/TA 60% (IQR 30-80%), caABMR 30% (IQR 10-40%), mixed rejection 40% (IQR 25-55%) and PCRR 40% (IQR 20-70%), overall *P*=0.001; **Supplementary Table 4 and 5**).

**Table 2.** – Semi-quantitative histological comparison of Banff scores for graft inflammation and graft fibrosis in the last included biopsy in different types of rejection.

| Banff score | ABMR<br>(N = 7) | TCMR<br>(N = 78) | Mixed<br>(N = 67) | PCRR<br>(N = 42) | p-value |
| --- | --- | --- | --- | --- | --- |
| **Graft inflammation** |  |  |  |  |  |
| t-score |  |  |  |  |  |
| i-score | 0 (0-0) | 2 (2-3) | 2 (1-3) | 3 (2-3) | < 0.001 |
| g-score | 0 (0-1) | 2 (1-3) | 3 (1-3) | 3 (1-3) | <0.001 |
| ti-score (%) | 2 (1-3) | 0 (0-0) | 2 (1-3) | 0 (0-1) | <0.001 |
| v-score | 30 (10-50) | 65 (30-90) | 80 (50-100) | 90 (70-100) | <0.001 |
| ptc-score | 0 (0-0) | 0 (0-1) | 1 (1-2) | 0 (0-0) | <0.001 |
|  | 3 (0-3) | 0 (0-0) | 3 (3-3) | 0.5 (0-3) | 0.000 |
| **Graft fibrosis** |  |  |  |  |  |
| IF/TA-score* | 20 (10-30) | 20 (10-30) | 10 (10-30) | 40 (20-70) | <0.001 |
| cg-score | 1 (0-3) | 0 (0-0) | 0 (0-2) | 0 (0-0) | <0.001 |
| cv-score | 1 (0-1.5) | 1 (0-2) | 1 (0-2) | 1 (0-1) | 0.88 |
| mm-score | 0 (0-1) | 0 (0-0) | 0 (0-0) | 0 (0-0) | <0.001 |
| ah-score | 1 (0-2) | 0 (0-1) | 0 (0-1) | 0 (0-1) | 0.01 |

\* IF/TA was scored semi-quantitative and expressed in %
Ordinal Banff scores are presented as median and interquartile range (IQR).
Abbreviations: TCMR = T-cell mediated rejection, ABMR = antibody-mediated rejection, PCRR = plasma cell rich rejection.
Banff scores: t = tubulitis, i = interstitial inflammation, g = glomerulitis, ti = total inflammation, v = intimal arteritis, ptc = peritubular capillaritis, IF/TA = interstitial fibrosis and tubular atrophy, cg = glomerular basement membrane (GBM) double contours, cv = vascular fibrous intimal thickening, mm = mesangial matrix expansion, ah = arteriolar hyalinosis.

### Rejection Class Algorithm

PCRR biopsies clusters mainly in cluster 3 and 6 based on acute components, which was most similar to TCMR and Mixed rejection (**Figure 4**). Cluster 3 is similar to acute TCMR in the Banff classification based on moderate to severe t- and i-scores. Cluster 6 contains biopsies which are HLA-DSA positive with high t- and i-score and microvascular inflammation and were shown to associated with highest risk of graft failure. Classification based on chronic components showed a more even distribution among all four categories, with highest percentages of biopsies in cluster 2 and especially cluster 3 compared to ABMR, TCMR and mixed rejection. Both clusters represent biopsies from a later stage of chronicity with longer time post-transplant. Cluster 3 is driven by high ct- and ci-scores while cluster 2 represents an intermediate phenotype.

**Figure 4.**
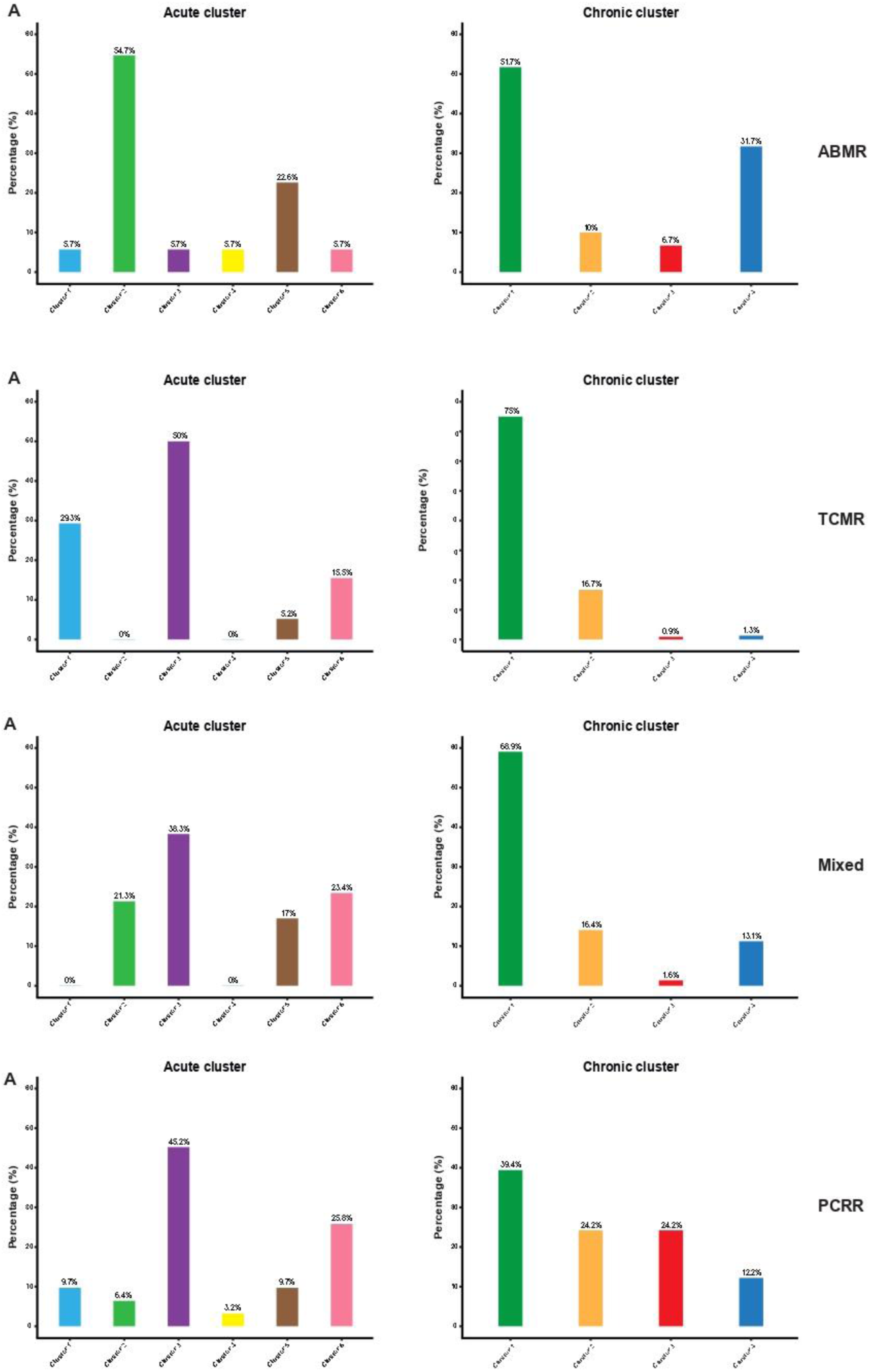
- Visualization of the distribution of the biopsies with different types of rejection among the different acute and chronic clusters based on the ‘RejectClass’ algorithm. Bar plots depicting the fractions of biopsies (as % of the total number of biopsies within that type of rejection) within each cluster based on acute lesions (acute clusters) and chronic lesions (chronic cluster). The classification among the clusters was generated using the algorithm on the website http://rejectionclass.eu.pythonanywhere.com and converted to bar plots visualizing (**A**) ABMR, (**B**) TCMR, (**C**) Mixed and (**D**) PCRR. The clusters are defined as described in the articles by Vaulet at al.^32–33^

### Clinical outcomes in PCRR compared to other types of rejection

**Figure 5A** displays Kaplan-Meier graft survival curves of PCRR compared to all types of rejection. Both PCRR and mixed rejection had similar poor graft survival but no overall significant differences (*P*=0.15). Compared to late types of rejection, no significant differences were observed (log-rank *P*=0.14; Figure 6A). PCRR showed a higher risk of graft loss compared to ABMR and TCMR with overall graft survival more similar to late types of rejection. After adjusting for fibrosis (IF/TA-score) and total inflammation (ti-score), no significant correlation between PCRR and increased risk of DCGF was observed (HR 0.86, 95% 0.46-1.61, *P=*0.64). A sensitivity analysis with a landmark set a one- and two-years post-transplant showed similar results (**Supplementary Figure 3 and 4**).

**Figure 5.**
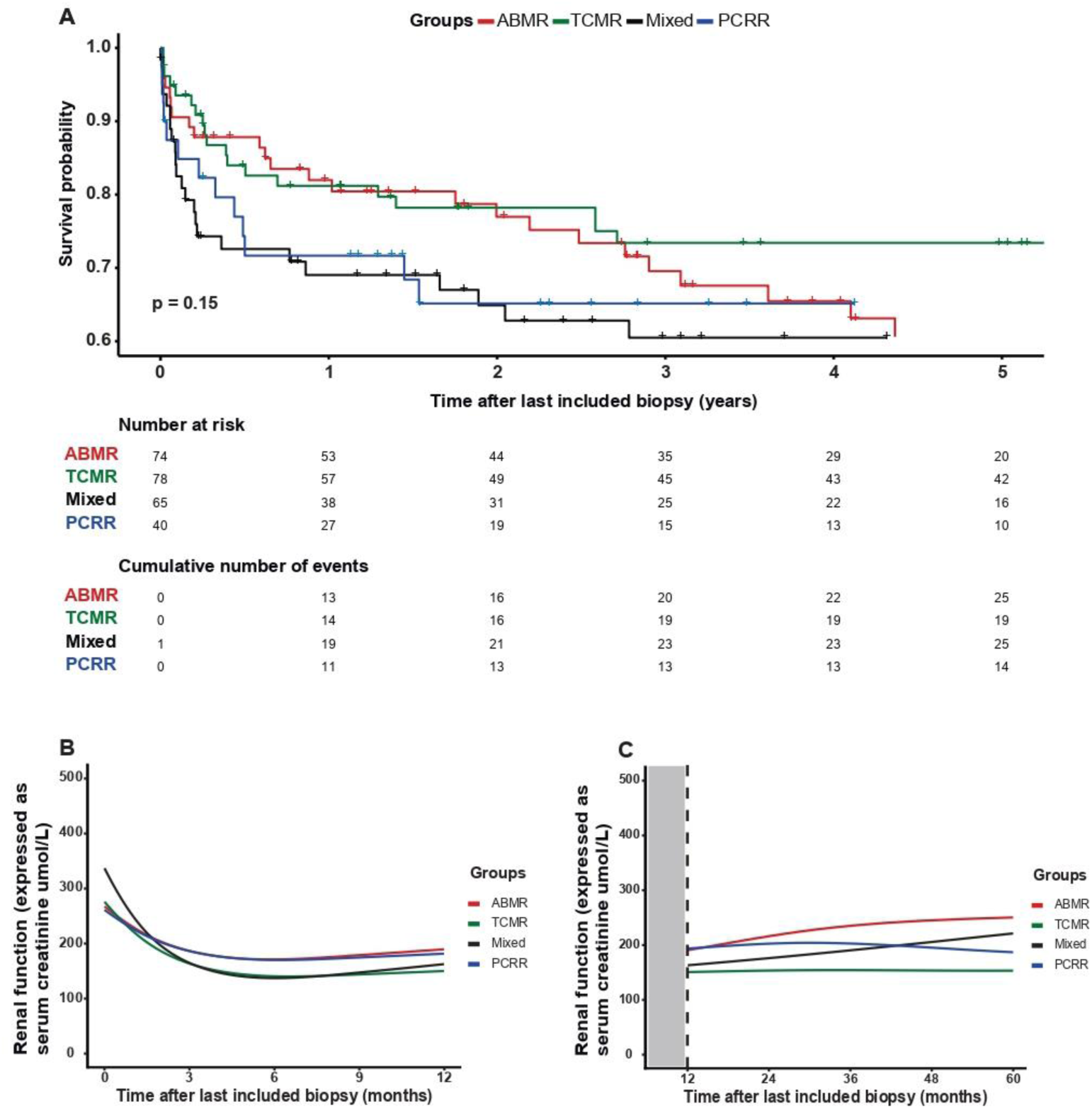
- Renal allograft outcome comparing PCRR to different types of rejection. (**A**) Kaplan Meier curves showing graft survival in patients with PCRR compared to all types of rejection (including acute, chronic-active and chronic ABMR, TCMR and mixed rejection) during a follow-up period of 5-year starting with the episode of rejection in the last included biopsy. (**B**) Linear mixed model analysis for renal function (expressed as serum creatinine levels in µmol/L) during the first 12 months after the last included biopsy, after adjusting for total inflammation (ti-score) and chronic damage (IF/TA-score). (**C**) Linear mixed model analysis with landmark analysis during year 1-5 after the last included biopsy after adjusting. Abbreviations: PCRR = plasma cell rich rejection, ABMR = antibody mediated rejection, TCMR = T-cell mediated rejection A visualization of the renal function showed the largest differences in the trajectory during the first year after the episode of rejection and stabilizing during the follow-up period after 1 year (**Supplementary Figure 5**). In a linear mixed model analysis, after adjusting for total inflammation (ti-score) and renal fibrosis (IF/TA-score), no significant differences were observed in slope of the renal function in PCRR compared to TCMR (β = 0.05, 95% CI = −0.18 – 0.27, *P*=0.69) or ABMR (β = 0.02, 95% CI = −0.23 – 0.27, *P*=0.86) within the first year after the episode of rejection (**Figure 5B**). In mixed rejection, renal

**Figure 6.**
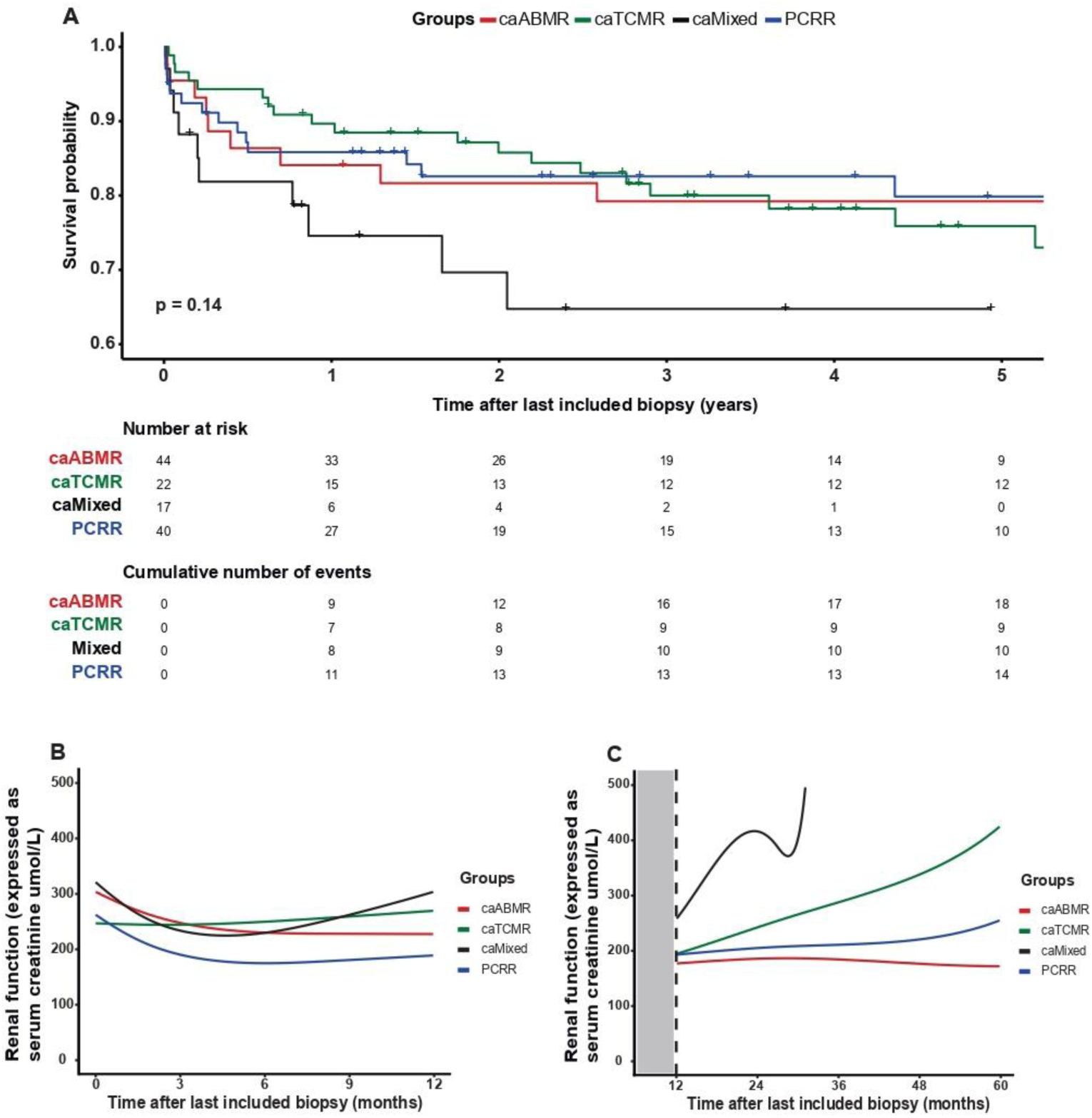
- Renal allograft outcome comparing PCRR to only late types of rejection. (**A**) Kaplan Meier curves showing graft survival in patients with PCRR compared to all types of rejection (including chronic-active and chronic ABMR, TCMR and mixed rejection) during a follow-up period of 5-year starting with the episode of rejection in the last included biopsy. (**B**) Linear mixed model analysis for renal function (expressed as serum creatinine levels in µmol/L) during the first 12 months after the last included biopsy, after adjusting for total inflammation (ti-score) and chronic damage (IF/TA-score). (**C**) Linear mixed model analysis with landmark analysis during year 1-5 after the last included biopsy after adjusting. function was significantly worse at the start of the follow-up period, but renal function improved quickly within 3 months after the start of the follow-up period (β = 0.24, 95% CI = 0.00 – 0.47, *P*=0.05). Renal function is similar within all four types of rejection > 1 year after the episode of rejection (**Figure 5C**). In the same analysis with only late rejections, no differences in renal function were observed within the first year after the episode of rejection, or > 1 year after (**Figure 6B and C**). In a joint model analysis, renal function was significantly associated with the risk of graft failure (β = 0.84, *P*=0.0001) with a time-dependent pattern in renal function. Only PCRR had a negative effect on renal function (β = −0.02, *P*=0.81 compared to ABMR, β = −0.12, *P*=0.10 to mixed). TCMR had significant positive effect (β = 0.15, *P*=0.03). None of the rejection types significantly affected the risk of graft failure directly.

### Previous episodes of rejection

In patients without any prior episodes of rejection (192/263 patients, 73%), the median interval from renal transplant to the diagnosis of their first episode of rejection in the last included biopsy was 25.2 months for PCRR (IQR 13-49 months), while the time interval until ABMR, TCMR or mixed rejection was significantly shorter (ABMR 11 months, IQR 0.5-49.2; TCMR 0.6 months, IQR 0.3-6.2 and mixed 0.4 months, IQR 0.3-1.9, overall *P*<0.001). In the PCRR group, 35/42 (83%) patients presented with PCRR as their initial presentation of rejection with the earliest presentation of PCRR 7 days after renal transplant in two patients. In both patients this was their first renal transplant, no pretransplant DSA were present and panel-reactive antibodies were low. The majority of patients (32/42, 76%) developed PCRR more than one year after renal transplant.

### Donor-specific antibodies

In patients who developed PCRR, 12/42 (29%) developed donor specific antibodies (DSA) post-transplant. Among these, 5/42 (12%) were *de novo* anti-HLA class I, 4/42 (10%) were *de novo* anti-HLA class II and 3/42 (7%) were *de novo* anti-HLA class I and II. There were no significant differences in the development of *de novo* anti-HLA antibodies between patients with PCRR and ABMR, TCMR or mixed rejection post-transplant (19/76 (25%) for ABMR; 12/78 (15%) for TCMR and 19/67 (28%) for mixed rejection, *P*=0.32). The mean survival time among patients with PCRR who developed *de novo* DSA post-transplant was 47±14 months and 117±19 months for patients with PCRR who did not develop *de novo* DSA post-transplant (*P*=0.32).

### Treatment in patient with PCRR

**Table 3** lists an overview of the immunosuppressive therapy in all 42 patients treated with PCRR. The admitted therapy in these patients consisted of medication currently used in either chronic/active TCMR, chronic/active ABMR of mixed rejection.

**Table 3.** – Overview of original diagnosis and subsequent treatment of the 42 patients classified as PCRR after revision of the biopsy.

| Patient number | Moment of biopsy after RT (months) | Original diagnosis | Treatment | DCGF* | Remarks |
| --- | --- | --- | --- | --- | --- |
| 1 | 3 | Mild tubulitis and interstitial nephritis with plasmacytosis consistent with ATN. No signs of acute rejection | No treatment | No |  |
| 2 | 0 | Acute TCMR I | Unknown | No |  |
| 3 | 5 | Acute TCMR III | MPNS | Yes |  |
| 4 | 11 | Acute TCMR III | MPNS and methylphenolic acid | Yes |  |
| 5 | 16 | Acute TCMR I with CNI toxicity | MPNS | No |  |
| 6 | 35 | Acute TCMR II | MPNS + ATG | No |  |
| 7 | 97 | Acute TCMR I with CNI toxicity | Unknown | No |  |
| 8 | 67 | CNI toxicity with early signs of CAG. No signs of active rejection. | No treatment | Yes |  |
| 9 | 0 | Acute TCMR II | ATG | No |  |
| 10 | 15 | Mixed rejection (Acute TCMR I and ABMR) | MPNS + ATG | Yes |  |
| 11 | 12 | Acute TCMR I | MPNS | No |  |
| 12 | 19 | Mixed rejection (Acute TCMR II and ABMR) | MPNS, plasmapheresis + ATG | Yes |  |
| 13 | 13 | Acute TCMR I | ATG | No |  |
| 14 | 50 | Acute ABMR with signs of CNI toxicity | Plasmapheresis and ATG | No |  |
| 15 | 23 | Chronic active TCMR | MPNS | No |  |
| 16 | 12 | Chronic active TCMR, Grade I | No treatment | No | Several opportunistic infections,<br>therefore no<br>treatment |
| 17 | 25 | Ongoing plasma-cell rich rejection | Unknown | No |  |
| 18 | 7 | Mixed rejection (plasma-cell rich active TCMR Ib and active ABMR) | No treatment | Yes | No possible treatment, back to hemodialysis |
| 19 | 85 | Plasma-cell rich rejection with minimal CAG | MPNS | Yes |  |
| 20 | 17 | Mixed rejection (chronic active TCMR and chronic active ABMR) | Plasmapheresis + ATG | No |  |
| 21 | 18 | Mixed rejection (Acute TCMR I and ABMR) | IVIg | No |  |
| 22 | 48 | Chronic active TCMR I | Plasmapheresis, MPNS, IVIg + Rituximab | No |  |
| 23 | 35 | Acute TCMR I, plasma-cell rich. Possible component active ABMR cannot be excluded based on C4d positivity | Plasmapheresis, MPNS + ATG | Yes |  |
| 24 | 12 | Borderline TCMR I | Unknown | No |  |
| 25 | 36 | Acute TCMR I, plasma-cell rich | MPNS | Yes |  |
| 26 | 37 | Acute TCMR II, plasma-cell rich | MPNS | Yes | No ATG due to severe chronic damage |
| 27 | 106 | Acute TCMR II, plasma-cell rich | MPNS | No |  |
| 28 | 142 | Chronic active ABMR | No treatment | Yes | Severe chronic damage, therefore no expected effect of treatment |
| 29 | 29 | Mixed rejection (Acute TCMR II, plasma-cell rich, and acute ABMR) | MPNS | No |  |
| 30 | 63 | Mixed rejection (chronic active TCMR and chronic active ABMR) | MPNS | No |  |
| 31 | 32 | Plasma-cell rich interstitial nephritis | No treatment | No |  |
| 32 | 21 | TMA, suspect for severe rejection | No treatment | No |  |
| 33 | 23 | Suspect for chronic active TCMR, Grade IA | MPNS | No |  |
| 34 | 70 | TMA with extensive lymphoplasmacellular infiltrate suspect for plasma-cell rich rejection | No treatment | Yes | Severe chronic damage, therefore no expected effect of treatment |
| 35 | 11 | Mixed rejection (Acute TCMR I and ABMR) | Plasmapheresis + IVIg | Yes |  |
| 36 | 46 | Mixed rejection (Acute TCMR II and ABMR) | Continue maintenance therapy (tacrolimus + MPNS) | No | No treatment possible due to severe psychiatric illness |
| 37 | 8 | Suspect for ABMR with early signs of vascular rejection, suspect for TCMR | Plasmapheresis + IVIG | No |  |
| 38 | 14 | Suspect for TCMR I | No treatment | No |  |
| 39 | 65 | Suspect for TCMR I | MPNS | No |  |
| 40 | 142 | Chronic active TCMR | Plasmapheresis, MPNS, IVIG + Rituximab | No |  |
| 41 | 76 | Mixed rejection (Acute TCMR, plasma-cell rich, and active ABMR) | Plasmapheresis, MPNS + ATG | No |  |
| 42 | 16 | Mixed rejection (chronic active TCMR and chronic active ABMR) | MPNS | Yes |  |

\* During a 5-year follow up period
Abbreviations: TCMR = T-cell mediated rejection, ABMR = antibody-mediated rejection, PCRR = plasma cell rich rejection, RT = renal transplant, ATN = acute tubulus necrosis, TMA = thrombotic microangiopathy, CNI = calcineurin inhibitor toxicity, ATG = antithymocyte immunoglobulin, IVIG = intravenous immunoglobulin, MPNS = methylprednisolone, DCGF = death-censored graft failure

## DISCUSSION

The primary objective of our study was to investigate the molecular characteristics of PCRR to gain insights into potential pathways involved in the pathogenesis of PCRR. Despite PCRR displaying more molecular heterogeneity, gene expression patterns differed from TCMR and ABMR suggesting involvement of distinct sets of genes and cell types in PCRR. PCRR typically manifests at a later stage after renal transplant and is associated with more pronounced graft inflammation and chronic damage. Our analysis showed that grafts survival and renal function in PCRR were more similar to late types of rejection, indicating close clinical follow-up of these patients is warranted due to higher risk of renal failure and graft loss.

Understanding the pathogenesis of PCRR and its underlying molecular mechanisms remains a challenge. One possible driving mechanism of PCRR could be a viral infection in association with tapering immunosuppressive therapy or non-adherence.^37^ Plasma cell infiltrates are often seen in association with BK nephropathy.^38,39^ In a case series by Hamada *et* al. all 6 patients had either preceding or concurrent viral infections, including BK, Varicella Zoster virus and Cytomegalovirus.^4^ In our study, we excluded cases with a concurrent viral infection during the current episode of rejection, but three patients within the group of PCRR did have a preceding BK nephropathy (more than 6 months before the biopsy). Another hypothesis centers on an exacerbated autoimmune response triggered by increased inflammation in the renal graft. This chronic stimulation and activation of B cells and maturation of plasma cells could be the driving factor of an autoimmune response. A recent report suggested some cases exhibit enrichment of innate-like B cells resembling mouse peritoneal B1 cells during rejection, which display broad auto-reactivity to inflamed renal tissues (e.g. autoantibodies against cytokines).^40^ Anecdotally, two patients from our center who developed graft failure with PCRR in their first transplant, experienced PCRR again in their second graft within 2 months and 4 years respectively, suggesting a potential memory B/plasma cell response. In both patients a concurrent viral infection was ruled out with immunohistochemical staining and additional blood tests. The hypothesis of a potential memory B/plasma cell response is also supported by the late development of PCRR in most patients (at least 3 months after renal transplant). To confirm whether PCRR is caused by an exacerbated autoimmune response, it is essential to identify the antigens B cells are exposed to and demonstrate the presence of local or circulating self- or alloantibodies. If PCRR is indeed driven by a memory B cell response, it could theoretically be associated with a higher risk of recurrence after the initial episode of PCRR. If these plasma cells are indeed causally related to graft failure, treatment with the new line of immunosuppressants targeting CD38 can be beneficial (Daratumumab, Felzartamab, Isatuximab).^41–43^ Alternatively, the influx of plasma cells in the graft could be a bystander effect reflective of prior damage without causal involvement in the pathogenesis leading to graft failure or, due to its morphological overlap with BKPyVAN, a presentation of a yet to discover viral infection.

One interesting finding was the enrichment of mast cells in biopsies with PCRR. Research has shown that mast cells play a role in various inflammatory kidney diseases, but also acute kidney injury and renal inflammation and fibrosis.^44–46^ The presence of mast cells in both acute and chronic renal allograft rejection has been described in previous studies, observing that an increased number of mast cells in explanted renal allografts with chronic rejection correlated to increased interstitial fibrosis.^47–49^ Microarray analysis revealed mast cell transcripts were associated with scarring in kidney transplants which in turn correlated with poor clinical prognosis.^31^ Our results also showed concomitant upregulation of *IL4*, *IL17F* and the *CPA3* enzyme involved in mast cell function. The presence of mast cells could potentially play an important role in the significant increase in graft inflammation and fibrosis observed in PCRR. Mast cells release mediators through degranulation (e.g., histamine), which induces vascular hyperpermeability and fluid extravasation, potentially explaining the presence of interstitial edema often observed in PCRR.^50^ Consequentially, mast cells may present a potential target for immunosuppressive treatment in patients with PCRR. Mast cell function can be directly inhibited by reducing their presence, modulating their activation or targeting downstream targets as has been described in cancer immunotherapy.^51^

Differences in graft survival and renal function in PCRR was not significant compared to other types of rejection. A large number of PCRR biopsies fitted into acute cluster 6 based on the Rejection Class algorithm, associated with highest risk of graft loss and presence of HLA-DSA. However, these clinical outcomes were poor in PCRR compared to ABMR, TCMR and mixed and their trajectory was more similar to late types of rejection. Overall graft survival of late types of rejection in general is unfavorable and patients with PCRR exhibited an equally poor graft survival as late type of rejections. Notably, the development of *de novo* DSA in patients with PCRR was associated with even worse graft survival than patients without DSA, a finding consistent with a study by Abbas et al.^5^ Our study further substantiates the notion that PCRR represents a late-onset form of rejection, supported by the results derived from the molecular data, particularly the pseudotime trajectory analysis. Additionally, we observed a significantly higher degree of chronic damage in biopsies with PCRR. Taken together, these results underscore that PCRR is indeed a late-stage event, substantiated by its molecular signature, the extent of chronic damage and time of diagnosis after renal transplant, especially when compared to (late) TCMR and ABMR. PCRR showed the highest level of chronicity using the Rejection Class algorithm. This observation could also explain the often poor response of PCRR to current treatment options, as higher degrees of chronic damage are indicative of a diminished treatment response.

The negative effect of PCRR on renal function and significant association between renal function and risk of graft failure highlights the critical importance of monitoring of renal function to for long-term allograft outcome. It highlights the need for early detection op PCRR, emphasizing the need for more tailored treatment of these patients and highlights the necessity for close communication with clinicians to facilitate intense clinical follow-up. Due to lack of targeted treatment of PCRR, most patients in our cohort received the standard immunosuppressive therapy for either (ca)TCMR and/or (ca)ABMR. Some case reports have reported the use of bortezomib as a possible treatment option for PCRR showing patients treated with bortezomib in conjunction to their standard immunosuppressive therapy, and moderate to severe IF/TA scores on biopsy, stabilized their renal function with 90% graft survival at 2-years after renal transplantation.^15^ Although these results seem promising, a better understanding of the pathways and cell types involved in PCRR are an important step towards the development of more targeted therapies.

A limitation of our study is the use of the B-HOT Nanostring panel, consisting of a limited panel of 770 genes based on consensus of recent literature in a 2019 Banff consensus meeting.^16^ However, selecting a panel of genes to be examined for mRNA analysis, inevitably introduces selection bias and limits the number of genes and pathways to be investigated. To obtain a more comprehensive understanding, methods involving more extensive transcriptional profiling should be used, for example RNA-sequencing. Another limitation of this study is the inclusion of only the last known biopsy with a biopsy-proven episode of rejection. The interval between renal transplant until the time of biopsy varied from several days up to 16 years (median 0.5 years). Aging of the kidney, prior episodes of rejection, viral infections, medication or other factors cause histopathological changes in the renal tissue, which are associated with molecular, structural and functional changes in the renal tissue. These changes elevate the risk of acute or chronic kidney disease.^52^ Therefore, it cannot be definitively concluded that all observed histological changes and degree of chronic damage present in the last biopsy is solely attributed to the episode of rejection present at the time of the biopsy; it may partly represent an accumulation of chronic damage over time.

Even though the exact mechanisms driving PCRR are yet to be completely eluded, to the best of our knowledge this study provides the first insights into the molecular and cellular composition of PCRR. PCRR is a late event post-transplant, both in terms of time of presentation post-transplant as well as the progression of chronic damage and molecular patterns. This study revealed that its molecular profile appears to be distinct from other types of rejection with clinical outcomes more similar to late types of rejection, and we therefore suggest PCRR should be considered as a separate entity in the next Banff classification. The question of whether PCRR is an exacerbated response to a viral infection or an auto-immune response of other entity remains uncertain underscoring the important need for more comprehensive research aimed at unravelling the underlying mechanism involved in PCRR. Such insights could enable a better and more tailored treatment for these patients and ultimately enhance long-term graft survival for patients affected by PCRR.

## Data Availability

All data produced in the present study are available upon reasonable request to the authors

## ACKNOWLEDGEMENTS

Dr. Florquin and Dr. Kers designed the study. Data collection and processing was performed by Ms. Chevalier Florquin and Ms. du Long. Practical experiments were performed by Ms. Chevalier Florquin and Mrs. Claessen. All biopsies were revised by dr. Florquin. The original draft of the manuscript was written by Ms. du Long. Statistical analysis was performed by Ms. du Long together with Dr. Peters-Sengers and Dr. Kers. The final version of the manuscript was revised and approved by all authors. This work was supported with funding from the Dutch Kidney Foundation grant no. DKF15OP09.

## Supplemental Methods

### mRNA gene expression analysis

Raw gene expression counts were imported and processed using the nSolver Analysis Software (Nanostring Technologies, version 4.0) and normalized counts were used for further analysis of differentially expressed genes. Significant differences in gene expression were identified using an adjusted p-value cutoff point of p < 0.05 and absolute log2foldchange >= 0.58 after log2 fold change (LFC) shrinkage. The adjusted p <0.05 was corrected for multiple testing using the Benjamini-Hochberg (BH) method to decrease the False Discovery Rate (FDR). We performed a principal component analysis (PCA) for linear dimensional reduction. The first 10 principal components (PCs) explained 88.6% of the total variances, with the first 2 PCs explaining 46.2% of the total variance visualized in a scree plot. Clustering classification among the three types of rejection (TCMR, ABMR and PCRR) was performed with t-distributed stochastic neighbor embedding (tSNE) for visualization of cluster classification. To examine whether PCRR has a different trajectory after renal transplant, and investigate if PCRR is a presentation of a similar or maybe different underlying diseases process as TCMR and ABMR, a pseudotime trajectory analysis was performed. With gene set enrichment analysis (GSEA) different cellular and biological processes enriched in PCRR, TCMR and ABMR were defined.

### Renal function

Renal function was based on serum creatinine levels. We tested six different linear mixed models using a different time moment as baseline measurement (T = 0) in each model: (1) serum creatinine level at the time of last included biopsy, (2) at time of renal transplant, (3) highest serum creatinine value measured during the month before last included biopsy, (4) highest serum creatinine value measured before the last included biopsy (between renal transplant and last included biopsy), (5) two weeks before last included biopsy and (6) one month before the of last included biopsy, Due to high number of missing values in model 5 and 6, we excluded these models for further analysis and continued the analysis with the other four models. The minimal adequate model for each analysis included type of rejection (‘rej_type1’), time, and their interaction (rej_type1*time) as fixed effects, and patient-specific intercept and slope of time as random effects. We fitted the mixed model (estimated using REML and optim optimizer) to predict the mean change in serum creatinine levels (‘Creat’) and compared the different models calculating the AIC (**Supplementary Figure 6**). The first model using the serum creatinine level at the time of the last included biopsy as baseline measurement for the follow-up period performed the best with the lowest AIC. This model was used for subsequent analysis adjusting for different confounders and landmark analysis. The 95% Confidence Intervals (CIs) and p-values were computed using a Wald t-distribution approximation. A two-sided p-value of < 0.05 was considered significant. The adjusted model included the following confounders: total inflammation (as expressed by the ti-scorre) and graft fibrosis (expressed by the IF/TA-score). Linearity of creatinine slope (applied to the variable time) was evaluated using a restricted cubic spline with 3 knots at default locations, and deemed nonlinear if the Wals statistic of the term was significant (*P*<0.05). Next, the same process was repeated for the mixed models analysis comparing PCRR to late rejection (**See Supplementary Figure 7**).

---

**Supplementary Table S1.** – Histopathological characteristics based on the Banff scores of the additional 24 biopsies used for the Nanostring mRNA analysis.

| Biopsy no. | Type of rejection | Graft inflammation |  |  |  |  |  | Graft fibrosis |  |  |  |  |
| --- | --- | --- | --- | --- | --- | --- | --- | --- | --- | --- | --- | --- |
|  |  | t-score | i-score | g-score | ti-score | v-score | ptc-score | IF/TA - score | cg-score | cv-score | mm-score | ah-score |
| 1 | ABMR | 0 | 1 | 2 | 50 | 1 | 3 | 20 | 0 | 1 | 0 | 1 |
| 2 | ABMR | 0 | 3 | 1 | 100 | 1 | 3 | 0 | 0 | 0 | 0 | 0 |
| 3 | ABMR | 3 | 3 | 3 | 70 | 2 | 3 | 20 | 0 | 1 | 0 | 2 |
| 4 | ABMR | 0 | 3 | 3 | 100 | 0 | 3 | 20 | 0 | 0 | 0 | 0 |
| 5 | ABMR | 0 | 0 | 3 | 30 | 0 | 3 | 30 | 0 | 1 | 0 | 0 |
| 6 | ABMR | 0 | 1 | 3 | 20 | 1 | 3 | 0 | 0 | 1 | 0 | 0 |
| 7 | ABMR | 0 | 1 | 3 | 10 | 1 | 3 | 0 | 0 | 1 | 0 | 0 |
| 8 | ABMR | 0 | 0 | 1 | 50 | 0 | 3 | 20 | 1 | 0 | 0 | 0 |
| 9 | PCRR | 3 | 3 | 0 | 100 | 2 | 0 | 20 | 0 | 3 | 0 | 0 |
| 10 | PCRR | 3 | 3 | 0 | 100 | 0 | 0 | 80 | 0 | 1 | 0 | 1 |
| 11 | PCRR | 3 | 2 | 0 | 50 | 0 | 3 | 70 | 0 | 1 | 0 | 1 |
| 12 | PCRR | 3 | 3 | 0 | 70 | 0 | 3 | 0 | 0 | 1 | 0 | 1 |
| 13 | PCRR | 3 | 3 | 0 | 100 | 0 | 3 | 90 | 0 | 2 | 0 | 0 |
| 14 | PCRR | 3 | 2 | 1 | 90 | 0 | 3 | 70 | 0 | 0 | 0 | 0 |
| 15 | PCRR | 1 | 1 | 3 | 30 | 0 | 3 | 10 | 0 | 1 | 0 | 0 |
| 16 | PCRR | 2 | 3 | 0 | 50 | 0 | 0 | 40 | 1 | 1 | 0 | 0 |
| 17 | TCMR | 3 | 13 | 0 | 40 | 0 | 0 | 20 | 0 | 2 | 0 | 0 |
| 18 | TCMR | 3 | 3 | 3 | 90 | 0 | 3 | 50 | 1 | 2 | 1 | 1 |
| 19 | TCMR | 3 | 3 | 1 | 50 | 1 | 0 | 0 | 0 | 2 | 0 | 0 |
| 20 | TCMR | 3 | 2 | 0 | 30 | 0 | 0 | 0 | 0 | 2 | 0 | 0 |
| 21 | TCMR | 2 | 3 | 0 | 70 | 0 | 0 | 0 | 0 | 0 | 0 | 1 |
| 22 | TCMR | 2 | 2 | 0 | 40 | 1 | 0 | 10 | 0 | 1 | 0 | 0 |
| 23 | TCMR | 3 | 3 | 0 | 70 | 1 | 0 | 10 | 0 | 1 | 0 | 0 |
| 24 | TCMR | 3 | 1 | 1 | 80 | 0 | 0 | 30 | 0 | 3 | 0 | 0 |

Abbreviations: TCMR = T-cell mediated rejection, ABMR = antibody-mediated rejection, PCRR = plasma cell rich rejection.
Banff scores: t = tubulitis, i = interstitial inflammation, g = glomerulitis, ti = total inflammation, v = intimal arteritis, ptc = peritubular capillaritis, IF/TA = interstitial fibrosis and tubular atrophy, cg = glomerular basement membrane (GBM) double contours, cv = vascular fibrous intimal thickening, mm = mesangial matrix expansion, ah = arteriolar hyalinosis

**Supplementary Table S2.**
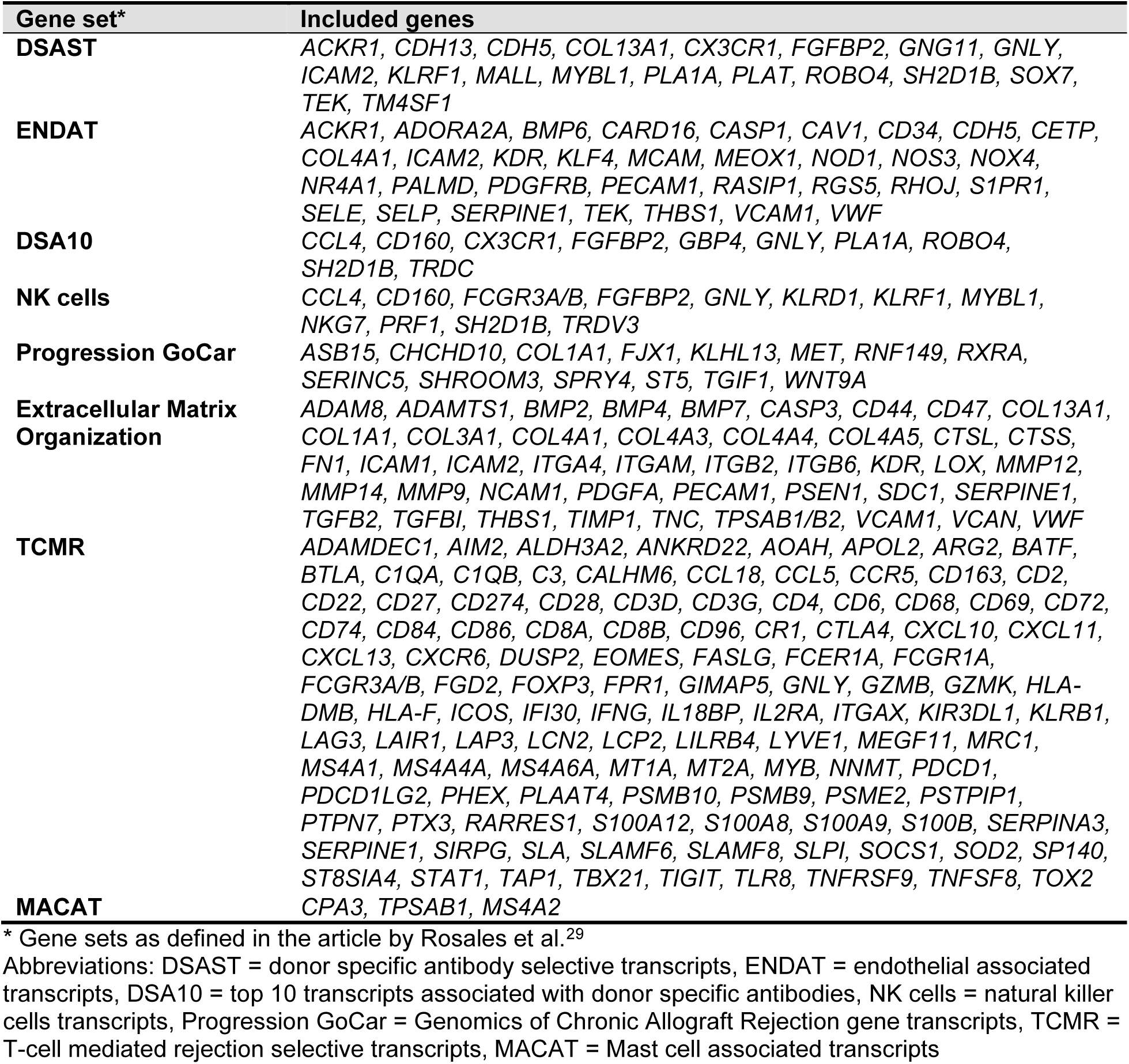
– Gene sets included in the Gene Set Enrichment Analysis (GSEA)

**Supplementary Table S3.** – Individual statistical comparison of Banff scores for graft inflammation and graft fibrosis of PCRR compared to the different types of rejection.

| Banff score | Overall p-value<sup>1</sup> | PCRR vs. ABMR<sup>2</sup> | PCRR vs. TCMR<sup>2</sup> | PCRR vs. Mixed<sup>2</sup> |
| --- | --- | --- | --- | --- |
| **Graft inflammation** |  |  |  |  |
| t-score | < 0.001 | < 0.001 | < 0.001 | < 0.001 |
| i-score | <0.001 | <0.001 | <0.001 | <0.001 |
| g-score | <0.001 | <0.001 | <0.001 | <0.001 |
| ti-score (%) | <0.001 | <0.001 | <0.001 | <0.001 |
| v-score | <0.001 | <0.001 | <0.001 | <0.001 |
| ptc-score | <0.001 | <0.001 | <0.001 | <0.001 |
| **Graft fibrosis** |  |  |  |  |
| IF/TA-score | <0.001 | < 0.001 | < 0.001 | < 0.001 |
| cg-score | <0.001 | < 0.001 | < 0.001 | < 0.001 |
| cv-score | 0.88 | 1.00 | 0.85 | 1.00 |
| mm-score | <0.001 | < 0.001 | < 0.001 | < 0.001 |
| ah-score | 0.01 | 0.01 | 0.65 | 0.41 |

<sup>1</sup> Kruskal Wallis H test
<sup>2</sup> Pairwise comparison using Dunn's procedure with a Bonferroni correction for multiple comparisons Abbreviations: TCMR = T-cell mediated rejection, ABMR = antibody-mediated rejection, PCRR = plasma cell rich rejection.
Banff scores: t = tubulitis, i = interstitial inflammation, g = glomerulitis, ti = total inflammation, v = intimal arteritis, ptc = peritubular capillaritis, IF/TA = interstitial fibrosis and tubular atrophy, cg = glomerular basement membrane (GBM) double contours, cv = vascular fibrous intimal thickening, mm = mesangial matrix expansion, ah = arteriolar hyalinosis

**Supplementary Table S4.** – Semi-quantitative histological comparison of Banff scores for graft inflammation and graft fibrosis in the last included biopsy comparing PCRR to late types of rejection.

| Banff score | caTCMR (N = 22) | caABMR (N = 45) | caMixed (N = 19) | PCRR (N = 42) | Overall p-value<sup>1</sup> |
| --- | --- | --- | --- | --- | --- |
| **Graft inflammation** |  |  |  |  |  |
| t-score |  |  |  |  |  |
| i-score | 2 (1-3) | 0 (0-0) | 2 (1-3) | 3 (2-3) | < 0.001 |
| g-score | 1 (0-2) | 0 (0-1) | 1 (1-2.5) | 3 (1-3) | <0.001 |
| ti-score | 0 (0-0) | 2 (2-3) | 3 (1.5-3) | 0 (0-1) | <0.001 |
| v-score | 70 (50-80) | 30 (20-50) | 70 (45-90) | 90 (70-100) | <0.001 |
| ptc-score | 0 (0-1) | 0 (0-0) | 1 (0-2) | 0 (0-0) | <0.001 |
|  | 0 (0-0) | 3 (0-3) | 3 (2.5-3) | 0.5 (0-3) | <0.001 |
| **Graft fibrosis** |  |  |  |  |  |
| IF/TA-score | 60 (30-80) | 30 (10-40) | 40 (25-55) | 40 (20-70) | 0.001 |
| cg-score | 0 (0-0) | 2 (2-3) | 2 (1-3) | 0 (0-0) | <0.001 |
| cv-score | 1.5 (1-3) | 1 (0-2) | 2 (1-3) | 1 (0-1) | <0.001 |
| mm-score | 0 (0-0) | 1 (0-1) | 1 (0-1) | 0 (0-0) | <0.001 |
| ah-score | 0 (0-1) | 1 (0-2) | 1 (0.5-2) | 0 (0-1) | <0.001 |

<sup>1</sup> Kruskal Wallis H test
Ordinal Banff scores are presented as median and interquartile range (IQR).
Abbreviations: caTCMR = chronic-active T-cell mediated rejection, caABMR = chronic-active antibody-mediated rejection, PCRR = plasma cell rich rejection. Banff scores: t = tubulitis, I = interstitial inflammation, g = glomerulitis, ti = total inflammation, v = intimal arteritis, ptc = peritubular capillaritis, IF/TA = interstitial fibrosis and tubular atrophy, cg = glomerular basement membrane (GBM) double contours, cv = vascular fibrous intimal thickening, mm = mesangial matrix expansion, ah = arteriolar hyalinosis

**Supplementary Table S5.** – Individual statistical comparison of Banff scores for graft inflammation and graft fibrosis of PCRR comparing PCRR to late types of rejection.

| Banff score | Overall p-value<sup>1</sup> | PCRR vs caABMR<sup>2</sup> | PCRR vs. caTBMR<sup>2</sup> | PCRR vs. caMixed<sup>2</sup> |
| --- | --- | --- | --- | --- |
| **Graft inflammation** |  |  |  |  |
| t-score | < 0.001 | < 0.001 | < 0.001 | < 0.001 |
| i-score | <0.001 | <0.001 | <0.001 | <0.001 |
| g-score | <0.001 | <0.001 | <0.001 | <0.001 |
| ti-score (%) | <0.001 | <0.001 | <0.001 | <0.001 |
| v-score | <0.001 | <0.001 | <0.001 | <0.001 |
| ptc-score | <0.001 | <0.001 | <0.001 | <0.001 |
| **Graft fibrosis** |  |  |  |  |
| IF/TA-score | 0.001 | 0.05 | 0.31 | 0.43 |
| cg-score | <0.001 | <0.001 | <0.001 | <0.001 |
| cv-score | <0.001 | 0.30 | 0.02 | <0.001 |
| mm-score | <0.001 | <0.001 | 0.81 | 0.001 |
| ah-score | <0.001 | 0.01 | 0.42 | 0.001 |

<sup>1</sup> Kruskal Wallis H test
<sup>2</sup> Pairwise comparison using Dunn's procedure with a Bonferroni correction for multiple comparisons Abbreviations: caTCMR = chronic-active T-cell mediated rejection, caABMR = chronic-active antibody-mediated rejection, PCRR = plasma cell rich rejection.
Banff scores: t = tubulitis, I = interstitial inflammation, g = glomerulitis, ti = total inflammation, v = intimal arteritis, ptc = peritubular capillaritis, IF/TA = interstitial fibrosis and tubular atrophy, cg = glomerular basement membrane (GBM) double contours, cv = vascular fibrous intimal thickening, mm = mesangial matrix expansion, ah = arteriolar hyalinosis

**Supplementary Figure S1.**
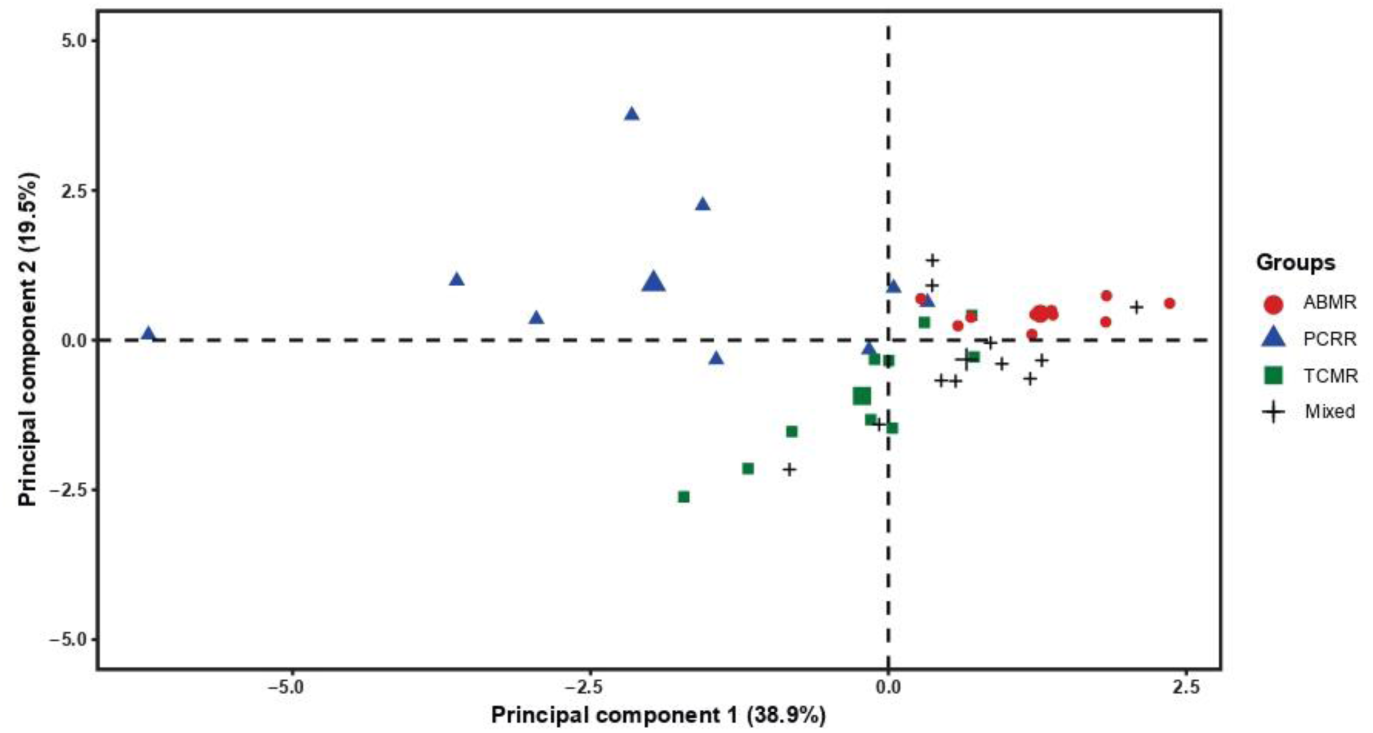
- Principal component analysis (PCA) Principal component analysis comparing expression of immunostainings in different types of rejections as a reflection of different cell types. The PCA plot visualizes immunohistochemical expression patterns comparing ABMR, PCRR, TCMR and mixed rejection including CD3, CD20, CD68, CD138, cKit and vimentin expression. Abbreviations: ABMR = antibody mediated rejection, TCMR = T-cell mediated rejection, PCRR = plasma cell rich rejection.

**Supplementary Figure S2.**
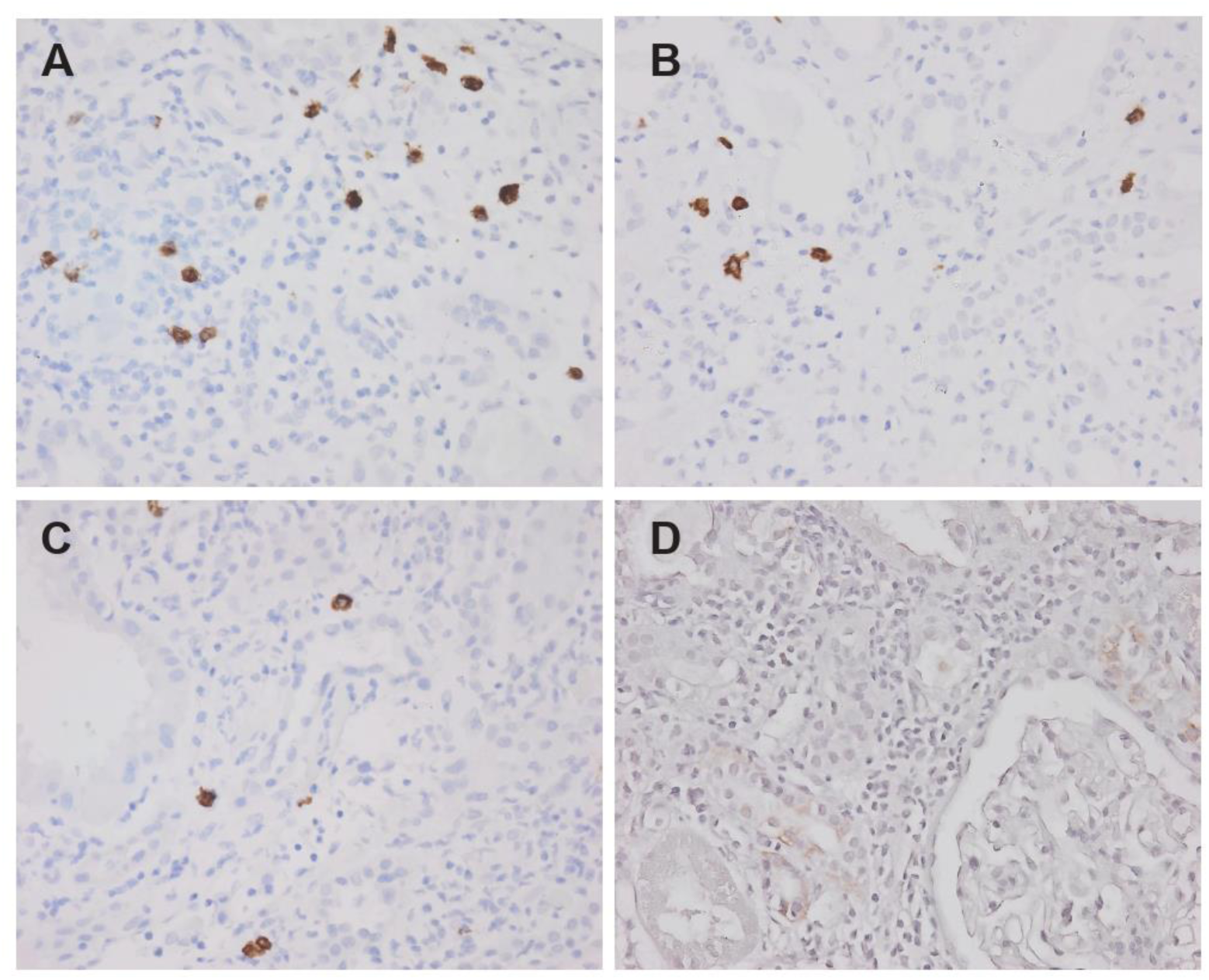
- Quantification of mast cells (based on the cKit staining) in different types of rejection. Example of the cKit staining (one high power field at 40x magnification) performed to assess and quantify the presence of mast cells in **(A)** PCRR, **(B)** TCMR, **(C)** ABMR and **(D)** mixed rejection. Abbreviations: ABMR = antibody mediated rejection, TCMR = T-cell mediated rejection, PCRR = plasma cell rich rejection.

**Supplementary Figure S3.**
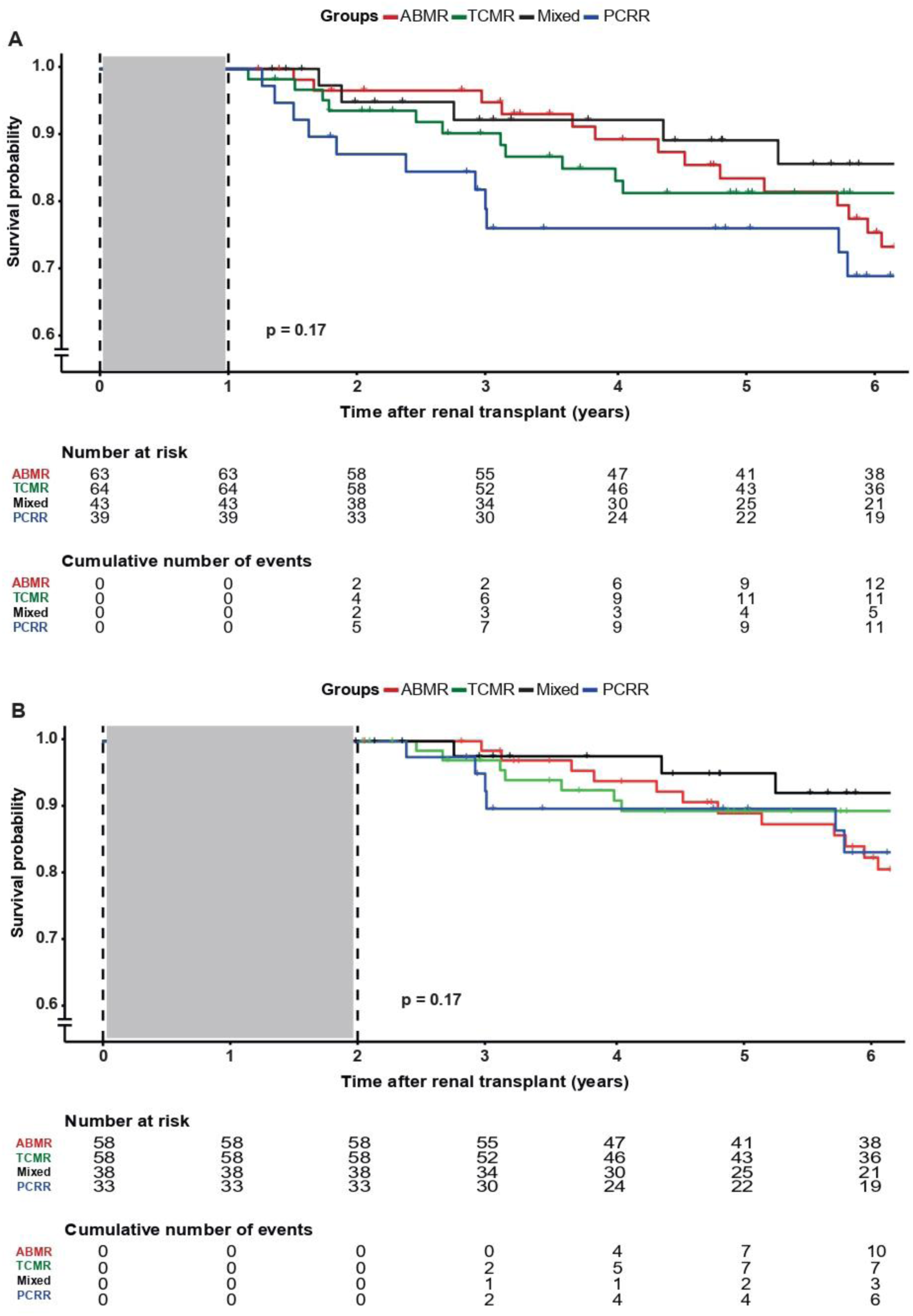
- Landmark survival analysis in PCRR versus all types of rejection. The upper panel **(A)** shows the graft survival in different types of rejection (including acute, chronic-active and chronic rejection) with a Kaplan Meier plot with a landmark set at one year post-transplant. The lower panel **(B)** shows the same graft survival analysis with a landmark set at two years post-transplant. Abbreviations: PCRR = plasma cell rich rejection, ABMR = antibody mediated rejection, TCMR = T-cell mediated rejection

**Supplementary Figure S4.**
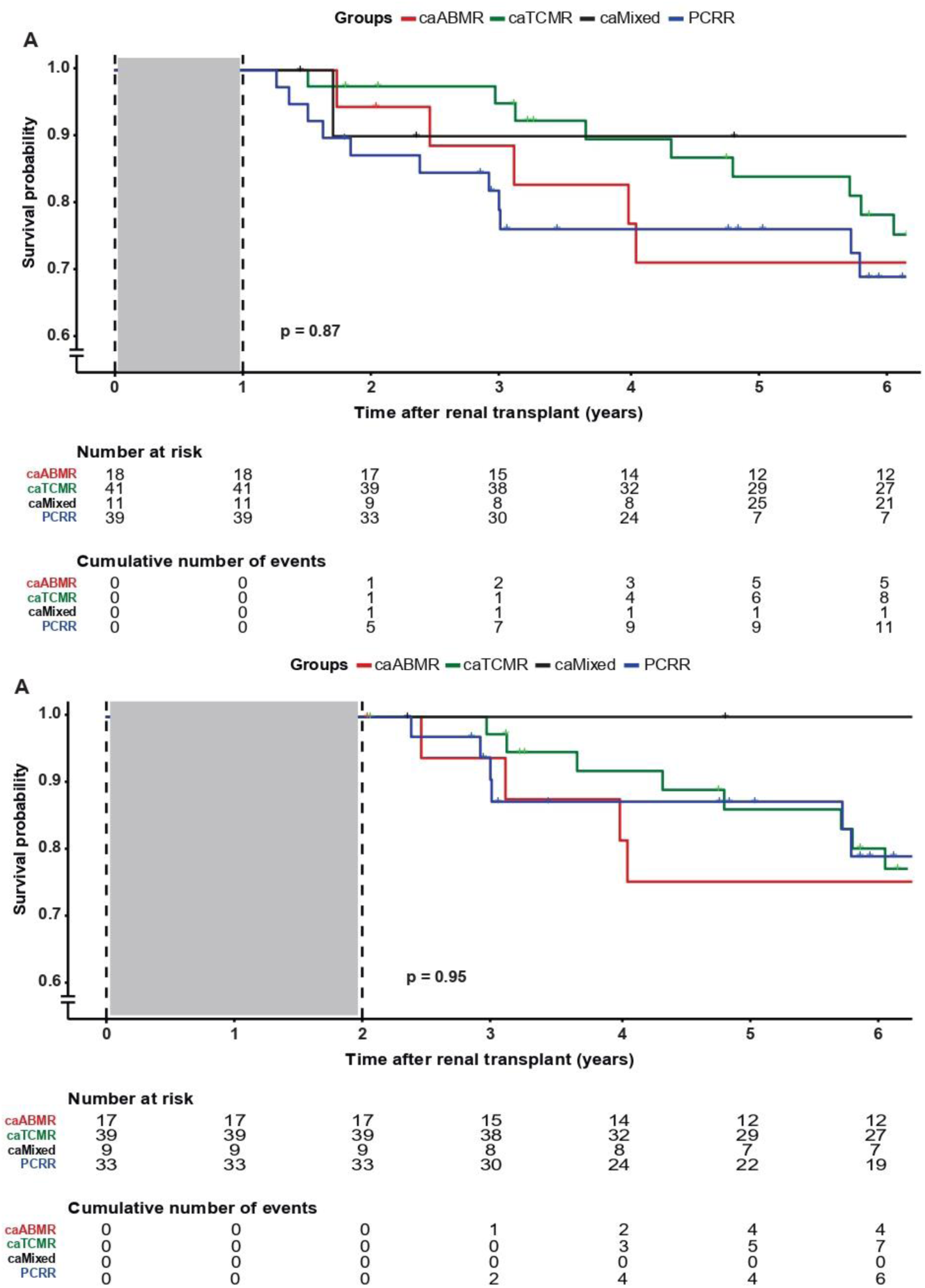
- Landmark survival analysis in PCRR versus late types of rejection. The upper panel **(A)** shows the graft survival in different types of rejection (including chronic-active and chronic rejection) with a Kaplan Meier plot with a landmark set at one year post-transplant. The lower panel **(B)** shows the same graft survival analysis with a landmark set at two years post-transplant. Abbreviations: PCRR = plasma cell rich rejection, caABMR = chronic-active antibody mediated rejection, caTCMR = chronic-active T-cell mediated rejection

**Supplementary Figure S5.**
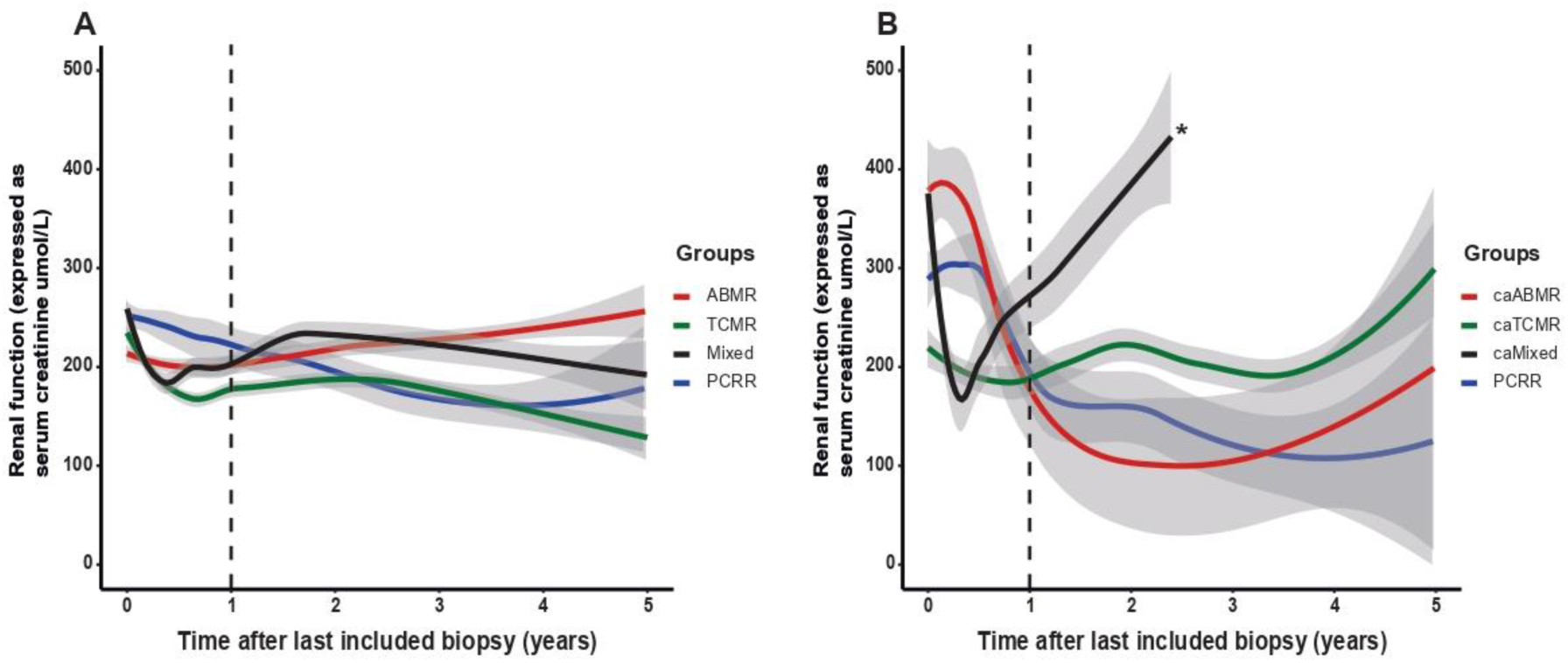
- Observed values of renal function during a 5-year follow-up period (A) Observed renal function (expressed as serum creatinine levels in µmol/L) in patients with ABMR, TCMR, mixed rejection and PCRR. The follow-up period is defined as a 5-year period starting at the moment of the last included biopsy (T = 0 on the x-axis). The largest differences in renal function between the four different types of rejection can be observed within the first year after the last included biopsy. A mixed model analysis was therefore performed to analyze renal function within the first year after the last biopsy and during year 1-5 after the last biopsy. **(B)** Observed renal function in patients with caABMR, caTCMR, caMixed rejection and PCRR during the same follow-up period. *All patients with caMixed rejection lost their graft due to death-censored graft failure within the first two year after the episode of caMixed rejection in the last included biopsy. Abbrevations: PCRR = plasma cell rich rejection, ABMR = antibody mediated rejection, TCMR = T-cell mediated rejection. caABMR = chronic-active ABMR, caTCMR = chronic-active TCMR, caMixed = chronic-active mixed rejection

**Supplementary Figure S6.**
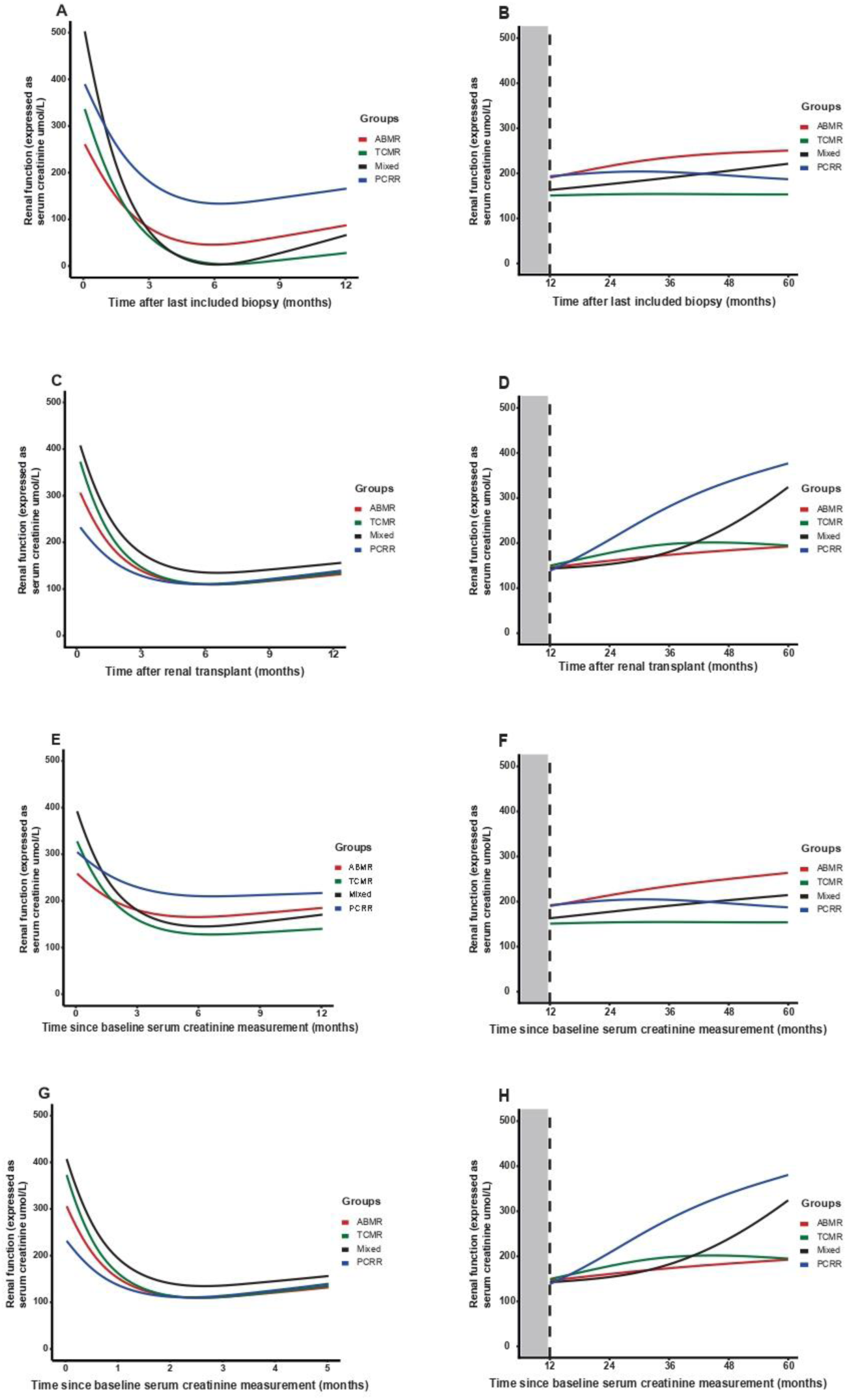
- Linear mixed model analysis using different models in all types of rejection. Linear mixed model analysis using a different moment around the time of the last included biopsy as baseline measurement (T = 0) including acute, chronic-active and chronic rejection. Renal function was evaluated at 0-12 months (first year) after the episode of rejection and 12-60 months (1-5 year). **(A-B)** Linear mixed model analysis with time of biopsy as T = 0 measurement, **(C-D)** time of renal transplant, **(E-F)** lowest serum creatinine level one month before last included biopsy and **(G-H)** lowest serum creatinine level between renal transplant and last biopsy. Analysis was performed using a minimal adequate unadjusted model. Abbreviations: PCRR = plasma cell rich rejection, ABMR = antibody mediated rejection, TCMR = T-cell mediated rejection

**Supplementary Figure S7.**
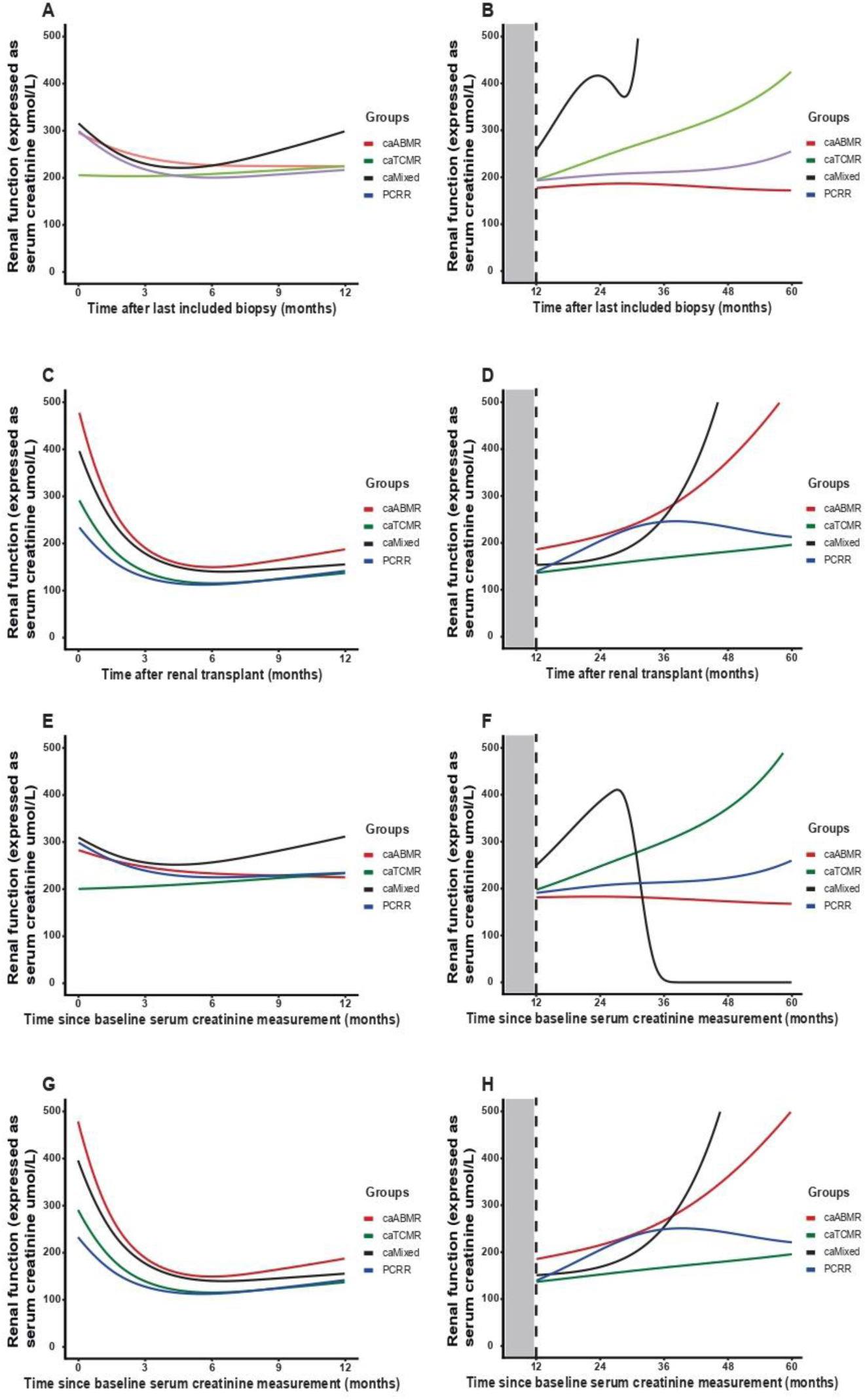
- Linear mixed model analysis using different models in late rejection. Linear mixed model analysis using a different moment around the time of the last included biopsy as baseline measurement (T = 0) including chronic-active and chronic rejection. Renal function was evaluated at 0-12 months (first year) after the episode of rejection and 12-60 months (1-5 year). **(A-B)** Linear mixed model analysis with time of biopsy as T = 0 measurement, **(C-D)** time of renal transplant, **(E-F)** lowest serum creatinine level one month before last included biopsy and **(G-H)** lowest serum creatinine level between renal transplant and last biopsy. Analysis was performed using a minimal adequate unadjusted model. Abbreviations: PCRR = plasma cell rich rejection, caABMR = chronic-active antibody mediated rejection, caTCMR = chronic-active T-cell mediated rejection

## References

1. Loupy A, Haas M, Roufosse C, et al. The Banff 2019 Kidney Meeting Report (I): Updates on and clarification of criteria for T cell- and antibody-mediated rejection. Am J Transplant. Sep 2020;20(9):2318-2331. doi:10.1111/ajt.15898

2. Demetris AJ, Bellamy C, Hübscher SG, et al. 2016 Comprehensive Update of the Banff Working Group on Liver Allograft Pathology: Introduction of Antibody-Mediated Rejection. Am J Transplant. Oct 2016;16(10):2816-2835. doi:10.1111/ajt.13909

3. Hasegawa J, Honda K, Omoto K, et al. Clinical and Pathological Features of Plasma Cell-Rich Acute Rejection After Kidney Transplantation. Transplantation. May 2018;102(5):853–859. doi:10.1097/tp.0000000000002041

4. Hamada AM, Yamamoto I, Kawabe M, et al. Clinicopathological features and outcomes of kidney allografts in plasma cell-rich acute rejection: A case series. Nephrology (Carlton). Jul 2018;23 Suppl 2:22–26. doi:10.1111/nep.13277

5. Abbas K, Mubarak M, Zafar MN, Aziz T, Abbas H, Muzaffar R, Rizvi SA. Plasma cell-rich acute rejections in living-related kidney transplantation: a clinicopathological study of 50 cases. Clin Transplant. Sep 2015;29(9):835–41. doi:10.1111/ctr.12589

6. Charney DA, Nadasdy T, Lo AW, Racusen LC. Plasma cell-rich acute renal allograft rejection. Transplantation. Sep 27 1999;68(6):791–7. doi:10.1097/00007890-199909270-00011

7. Nishimura A, Masuzawa N, Nakamura T, et al. Clinicopathological and immunohistochemical analysis of plasma cell-rich rejection in renal transplantation: Involvement of intratubular Th1/Th2 balance in plasma cell enrichment. Nephrology (Carlton). Jul 2018;23 Suppl 2:52–57. doi:10.1111/nep.13273

8. Desvaux D, Le Gouvello S, Pastural M, et al. Acute renal allograft rejections with major interstitial oedema and plasma cell-rich infiltrates: high gamma-interferon expression and poor clinical outcome. Nephrol Dial Transplant. Apr 2004;19(4):933–9. doi:10.1093/ndt/gfh027

9. Plaza Lara E, Hernández García E, Ruiz Fuentes MDC, Caba Molina M, De Gracia Guindo MDC, Osuna Ortega A. Plasma Cell-Rich Acute Rejection in Renal Transplantation: A Case Report. Transplant Proc. Mar 2020;52(2):512–514. doi:10.1016/j.transproceed.2019.12.015

10. Chang A, Moore JM, Cowan ML, et al. Plasma cell densities and glomerular filtration rates predict renal allograft outcomes following acute rejection. Transpl Int. Oct 2012;25(10):1050–8. doi:10.1111/j.1432-2277.2012.01531.x

11. Gärtner V, Eigentler TK, Viebahn R. Plasma cell-rich rejection processes in renal transplantation: morphology and prognostic relevance. Transplantation. Apr 15 2006;81(7):986–91. doi:10.1097/01.tp.0000215014.40595.ab

12. Carpio VN, Noronha Ide L, Martins HL, et al. Expression patterns of B cells in acute kidney transplant rejection. Exp Clin Transplant. Oct 2014;12(5):405–14.

13. Mubarak M, Abbas K, Zafar MN, Aziz T. Immunohistopathologic Characterization of Plasma Cell-Rich Acute Rejection in Living-Related Renal Transplant Recipients. Exp Clin Transplant. Oct 2017;15(5):516–520. doi:10.6002/ect.2016.0188

14. Kers J, Bülow RD, Klinkhammer BM, et al. Deep learning-based classification of kidney transplant pathology: a retrospective, multicentre, proof-of-concept study. Lancet Digit Health. Jan 2022;4(1):e18–e26. doi:10.1016/s2589-7500(21)00211-9

15. Abbas K, Mubarak M, Zafar MN, Musharraf W, Imam M, Aziz T, Rizvi AH. Management of Plasma Cell-Rich Acute Rejection in Living-Related Kidney Transplant: Role of Proteasome Inhibitor. Exp Clin Transplant. Feb 2019;17(1):42–46. doi:10.6002/ect.2017.0154

16. Mengel M, Loupy A, Haas M, et al. Banff 2019 Meeting Report: Molecular diagnostics in solid organ transplantation-Consensus for the Banff Human Organ Transplant (B-HOT) gene panel and open source multicenter validation. Am J Transplant. Sep 2020;20(9):2305-2317. doi:10.1111/ajt.16059

17. Varol H, Ernst A, Cristoferi I, et al. Feasibility and Potential of Transcriptomic Analysis Using the NanoString nCounter Technology to Aid the Classification of Rejection in Kidney Transplant Biopsies. Transplantation. Apr 1 2023;107(4):903–912. doi:10.1097/tp.0000000000004372

18. Smith RN, Rosales IA, Tomaszewski KT, et al. Utility of Banff Human Organ Transplant Gene Panel in Human Kidney Transplant Biopsies. Transplantation. May 1 2023;107(5):1188–1199. doi:10.1097/tp.0000000000004389

19. Newman AM, Liu CL, Green MR, et al. Robust enumeration of cell subsets from tissue expression profiles. Nat Methods. May 2015;12(5):453–7. doi:10.1038/nmeth.3337

20. Halloran PF, Madill-Thomsen KS, Reeve J. The Molecular Phenotype of Kidney Transplants: Insights From the MMDx Project. Transplantation. Jan 1 2024;108(1):45–71. doi:10.1097/tp.0000000000004624

21. Hidalgo LG, Sis B, Sellares J, et al. NK cell transcripts and NK cells in kidney biopsies from patients with donor-specific antibodies: evidence for NK cell involvement in antibody-mediated rejection. Am J Transplant. Aug 2010;10(8):1812–22. doi:10.1111/j.1600-6143.2010.03201.x

22. Sis B, Jhangri GS, Bunnag S, Allanach K, Kaplan B, Halloran PF. Endothelial gene expression in kidney transplants with alloantibody indicates antibody-mediated damage despite lack of C4d staining. Am J Transplant. Oct 2009;9(10):2312–23. doi:10.1111/j.1600-6143.2009.02761.x

23. Halloran PF, Matas A, Kasiske BL, Madill-Thomsen KS, Mackova M, Famulski KS. Molecular phenotype of kidney transplant indication biopsies with inflammation in scarred areas. Am J Transplant. May 2019;19(5):1356–1370. doi:10.1111/ajt.15178

24. Halloran PF, Venner JM, Famulski KS. Comprehensive Analysis of Transcript Changes Associated With Allograft Rejection: Combining Universal and Selective Features. Am J Transplant. Jul 2017;17(7):1754–1769. doi:10.1111/ajt.14200

25. Halloran PF, Venner JM, Madill-Thomsen KS, Einecke G, Parkes MD, Hidalgo LG, Famulski KS. Review: The transcripts associated with organ allograft rejection. Am J Transplant. Apr 2018;18(4):785–795. doi:10.1111/ajt.14600

26. Madill-Thomsen KS, Böhmig GA, Bromberg J, et al. Donor-Specific Antibody Is Associated with Increased Expression of Rejection Transcripts in Renal Transplant Biopsies Classified as No Rejection. J Am Soc Nephrol. Nov 2021;32(11):2743–2758. doi:10.1681/asn.2021040433

27. Venner JM, Hidalgo LG, Famulski KS, Chang J, Halloran PF. The molecular landscape of antibody-mediated kidney transplant rejection: evidence for NK involvement through CD16a Fc receptors. Am J Transplant. May 2015;15(5):1336–48. doi:10.1111/ajt.13115

28. O’Connell PJ, Zhang W, Menon MC, et al. Biopsy transcriptome expression profiling to identify kidney transplants at risk of chronic injury: a multicentre, prospective study. Lancet. Sep 3 2016;388(10048):983–93. doi:10.1016/s0140-6736(16)30826-1

29. Menon MC, Chuang PY, Li Z, et al. Intronic locus determines SHROOM3 expression and potentiates renal allograft fibrosis. J Clin Invest. Jan 2015;125(1):208–21. doi:10.1172/jci76902

30. Rosales IA, Mahowald GK, Tomaszewski K, et al. Banff Human Organ Transplant Transcripts Correlate with Renal Allograft Pathology and Outcome: Importance of Capillaritis and Subpathologic Rejection. J Am Soc Nephrol. Dec 2022;33(12):2306–2319. doi:10.1681/asn.2022040444

31. Mengel M, Reeve J, Bunnag S, et al. Molecular correlates of scarring in kidney transplants: the emergence of mast cell transcripts. Am J Transplant. Jan 2009;9(1):169–78. doi:10.1111/j.1600-6143.2008.02462.x

32. Vaulet T, Divard G, Thaunat O, et al. Data-Driven Chronic Allograft Phenotypes: A Novel and Validated Complement for Histologic Assessment of Kidney Transplant Biopsies. J Am Soc Nephrol. Nov 2022;33(11):2026–2039. doi:10.1681/asn.2022030290

33. Vaulet T, Divard G, Thaunat O, et al. Data-driven Derivation and Validation of Novel Phenotypes for Acute Kidney Transplant Rejection using Semi-supervised Clustering. J Am Soc Nephrol. May 3 2021;32(5):1084–1096. doi:10.1681/asn.2020101418

34. Lai X, Zheng X, Mathew JM, Gallon L, Leventhal JR, Zhang ZJ. Tackling Chronic Kidney Transplant Rejection: Challenges and Promises. Front Immunol. 2021;12:661643. doi:10.3389/fimmu.2021.661643

35. Rizopoulos D. The R Package JMbayes for Fitting Joint Models for Longitudinal and Time-to-Event Data Using MCMC. Journal of Statistical Software. 08/28 2016;72(7):1 - 46. doi:10.18637/jss.v072.i07

36. Gleiss A, Oberbauer R, Heinze G. An unjustified benefit: immortal time bias in the analysis of time-dependent events. Transpl Int. Feb 2018;31(2):125–130. doi:10.1111/tri.13081

37. Hasegawa J, Honda K, Wakai S, et al. Plasma Cell-Rich Rejection After Kidney Transplantation and the Role of Donor-Specific Antibodies: A Case Report and Review of the Literature. Transplant Proc. Oct 2015;47(8):2533–6. doi:10.1016/j.transproceed.2015.09.018

38. Kemény E, Hirsch HH, Eller J, Dürmüller U, Hopfer H, Mihatsch MJ. Plasma cell infiltrates in polyomavirus nephropathy. Transpl Int. Apr 1 2010;23(4):397–406. doi:10.1111/j.1432-2277.2009.01001.x

39. Buettner M, Xu H, Böhme R, et al. Predominance of TH2 cells and plasma cells in polyoma virus nephropathy: a role for humoral immunity? Hum Pathol. Sep 2012;43(9):1453–62. doi:10.1016/j.humpath.2011.11.006

40. Asano Y, Daccache J, Jain D, et al. Innate-like self-reactive B cells infiltrate human renal allografts during transplant rejection. Nat Commun. Jul 16 2021;12(1):4372. doi:10.1038/s41467-021-24615-6

41. Vincenti F, Bestard O, Brar A, et al. Isatuximab Monotherapy for Desensitization in Highly Sensitized Patients Awaiting Kidney Transplant. J Am Soc Nephrol. Dec 26 2023;doi:10.1681/asn.0000000000000287

42. Doberer K, Kläger J, Gualdoni GA, et al. CD38 Antibody Daratumumab for the Treatment of Chronic Active Antibody-mediated Kidney Allograft Rejection. Transplantation. Feb 1 2021;105(2):451–457. doi:10.1097/tp.0000000000003247

43. Mayer KA, Budde K, Halloran PF, et al. Safety, tolerability, and efficacy of monoclonal CD38 antibody felzartamab in late antibody-mediated renal allograft rejection: study protocol for a phase 2 trial. Trials. Apr 8 2022;23(1):270. doi:10.1186/s13063-022-06198-9

44. Mack M, Rosenkranz AR. Basophils and mast cells in renal injury. Kidney Int. Dec 2009;76(11):1142–7. doi:10.1038/ki.2009.320

45. Madjene LC, Pons M, Danelli L, et al. Mast cells in renal inflammation and fibrosis: lessons learnt from animal studies. Mol Immunol. Jan 2015;63(1):86–93. doi:10.1016/j.molimm.2014.03.002

46. Owens EP, Vesey DA, Kassianos AJ, Healy H, Hoy WE, Gobe GC. Biomarkers and the role of mast cells as facilitators of inflammation and fibrosis in chronic kidney disease. Transl Androl Urol. May 2019;8(Suppl 2):S175–s183. doi:10.21037/tau.2018.11.03

47. Danilewicz M, Wagrowska-Danilewicz M. Immunohistochemical analysis of the interstitial mast cells in acute rejection of human renal allografts. Med Sci Monit. May 2004;10(5):Br151-6.

48. Yamada M, Ueda M, Naruko T, et al. Mast cell chymase expression and mast cell phenotypes in human rejected kidneys. Kidney Int. Apr 2001;59(4):1374–81. doi:10.1046/j.1523-1755.2001.0590041374.x

49. Pardo J, Diaz L, Errasti P, et al. Mast cells in chronic rejection of human renal allografts. Virchows Arch. Aug 2000;437(2):167–72. doi:10.1007/s004280000211

50. Nguyen SMT, Rupprecht CP, Haque A, Pattanaik D, Yusin J, Krishnaswamy G. Mechanisms Governing Anaphylaxis: Inflammatory Cells, Mediators, Endothelial Gap Junctions and Beyond. Int J Mol Sci. Jul 21 2021;22(15)doi:10.3390/ijms22157785

51. Lichterman JN, Reddy SM. Mast Cells: A New Frontier for Cancer Immunotherapy. Cells. May 21 2021;10(6)doi:10.3390/cells10061270

52. Fang Y, Gong AY, Haller ST, Dworkin LD, Liu Z, Gong R. The ageing kidney: Molecular mechanisms and clinical implications. Ageing Res Rev. Nov 2020;63:101151. doi:10.1016/j.arr.2020.101151

